# Defining and emulating target trials of the effects of postexposure vaccination using observational data

**DOI:** 10.1101/2023.05.03.23289471

**Authors:** Christopher Boyer, Marc Lipsitch

## Abstract

Postexposure vaccination has the potential to prevent or modify the course of clinical disease among those exposed to a pathogen. However, due to logistical constraints, postexposure vaccine trials have been difficult to implement in practice. In place of trials, investigators have used observational data to estimate the effectiveness or optimal timing window for postexposure vaccines, but the relationship between these analyses and those that would be conducted in a trial is often unclear. Here, we define several possible target trials for postexposure vaccination and show how, under certain conditions, they can be emulated using observational data. We emphasize the importance of the incubation period and the timing of vaccination in trial design and emulation. As an example, we specify a protocol for postexposure vaccination against mpox and provide a step-by-step description of how to emulate it using data from a healthcare database or contact tracing program. We further illustrate some of the benefits of the target trial approach through simulation.

## 1 Introduction

For a millennium or more humans have been inoculating healthy, unexposed individuals to prevent the onset of future disease [1]. Today, this remains the dominant paradigm for the development and mass administration of vaccines. By contrast, using vaccines to prevent clinical disease among those *already exposed* to a pathogen, i.e. postexposure vaccination, remains an under-utilized strategy despite its potential to curb outbreaks and prevent the worst sequelae of disease [2]. This is due, in part, to the difficulty of running postexposure trials, particularly during a large outbreak, as investigators must identify, randomize, and vaccinate participants all in the, often short, time window between exposure and symptom onset. Even when these trials are feasible, the effectiveness of a postexposure vaccine is likely to vary dramatically based on the time elapsed since exposure, which can make it difficult to compare estimates across trials with different distributions of vaccination times. Finally, when there is other evidence to support effectiveness, and when other treatments are unavailable, a randomized postexposure trial may be considered unethical.

In the absence of trial data, an alternative approach is to use observational data to emulate the trial desired [3, 4] (called a “target trial”), for instance by using electronic healthcare records or public health contact tracing databases to define cohorts of individuals exposed to infection and comparing outcomes among those who do and do not receive post-exposure vaccination. In this paper, we define several target trials for assessing the effectiveness of postexposure vaccination depending on the causal quantity of interest (Note, in a slight abuse of terminology we refer to vaccine effectiveness rather than efficacy throughout, even when the target trial itself could reasonably be called an efficacy trial because, ultimately, the observational data used for the emulation are collected under real world conditions). We also discuss the conditions under which a trial can be emulated from observational data. We show how the target trial framework can help clarify the causal question and resolve common biases in the observational analysis of postexposure effectiveness through alignment of time zero, eligibility, and assignment as well as an unambiguous definition of the treatment strategies being contrasted. We provide an example protocol for emulating a trial of a postexposure vaccine for mpox and illustrate some of the benefits of this approach through simulation. Throughout, we focus on the direct effect of vaccination [5] on the individuals receiving a postexposure vaccine although the approach could be generalized to include spillover effects on others in the population.

## 2 Design challenges: incubation period and timing of vaccination

The success of postexposure prophylactic vaccination is determined by two competing forces: the incubation period of the pathogen and how long it takes to receive a vaccine. To provide benefit postexposure, a vaccine must stimulate an immune response faster, greater, or more specific than that provoked by natural infection alone. For example, in the case of smallpox, a vaccine administered within 72 hours after exposure to the variola virus (the causative virus of smallpox) induces an antibody response 4 to 8 days earlier than the variola virus itself, most likely because the vaccine response bypasses the initial stages of natural infection in the respiratory tract [6, 7]. However, delays in receiving a vaccine are common as participants must first either recognize or be notified of their exposure and then present at a healthcare clinic where a vaccine is available.

The resulting overlap between the timing of vaccination and the timing of symptom onset creates several design challenges (see Figure 1). First, the effectiveness of a vaccine may vary substantially depending on how quickly participants can be vaccinated postexposure (top panel, Figure 1). In a randomized trial, the protocol specifies the precise vaccination strategy to be evaluated and must strike a balance between existing exposure identification, enrollment, and care coordination systems and what is known about the biology governing the natural course of infection. This can be difficult when the incubation period or mechanism of action of a postexposure vaccine are not well established. Under these circumstances, longer delays may be permitted with a secondary goal to infer the optimal postexposure window to administer the vaccine. In an observational setting, by contrast, the protocol for vaccine timing is often less clear or may even be absent, in which case the vaccination strategy being evaluated may be ambiguous.

**Figure 1.**
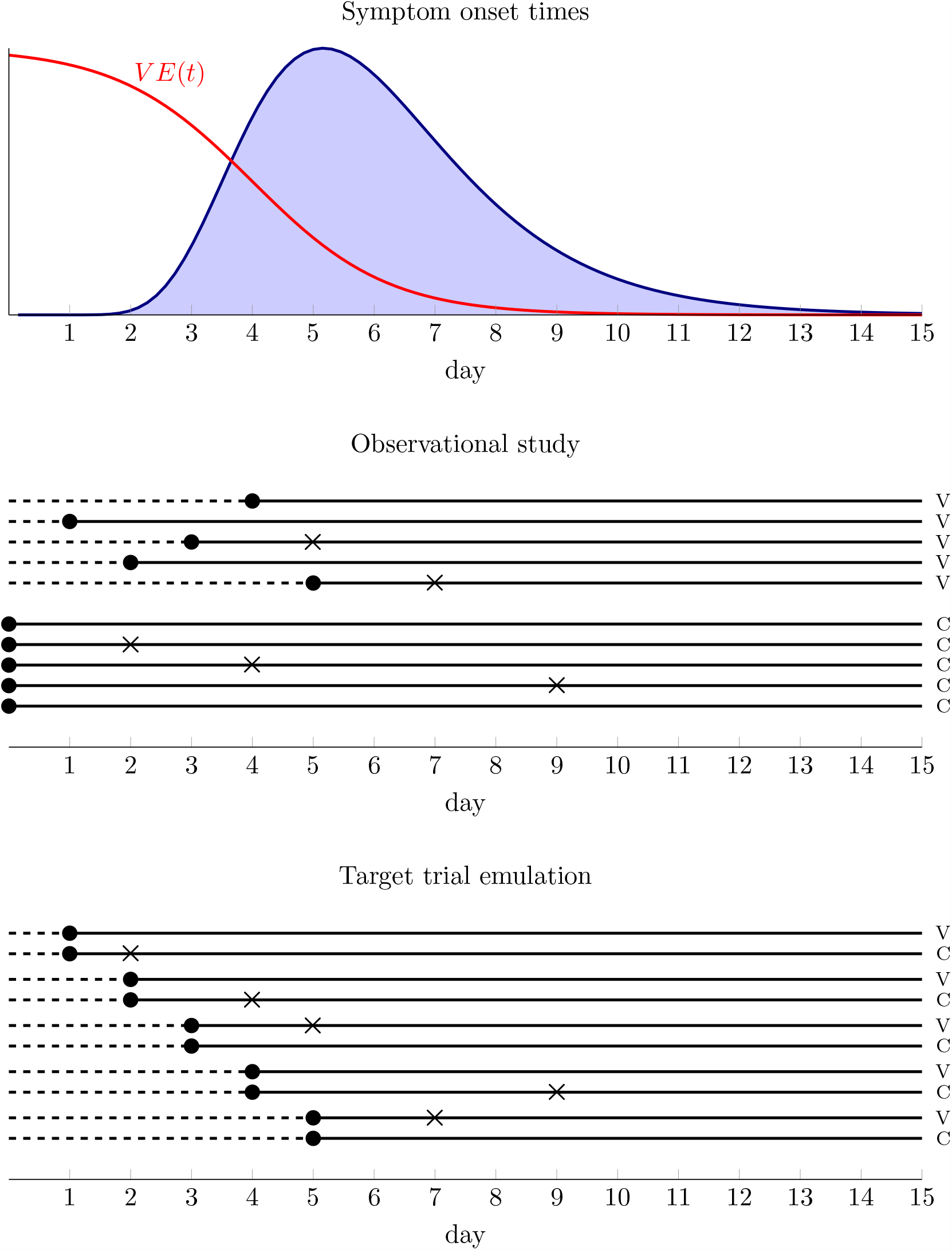
Illustration of the challenges of evaluating postexposure vaccination using observational data. The top panel shows the distribution of symptom onset times among cases as well as vaccine effectiveness as a function of postexposure day of administration for a hypothetical pathogen. The middle panel shows an observational study with 5 vaccinated (V) and 5 unvaccinated (C) individuals in which there are delays in receiving vaccines. Dots show the time exposure status is first defined and Xs show symptom onset. The dashed line represents possible immortal time among vaccinated who have to survive symptom free long enough to be vaccinated. The bottom panel shows a nested sequence of daily trials among the same individuals in which there is no immortal time bias because the timing of enrollment and exposure assignment coincides in each trial.

Second, when vaccination is delayed there is also the possibility that some participants may have already developed symptoms prior to enrollment or vaccination. A vaccine can prevent symptoms only if administered before those symptoms start. However, when those who have symptoms at enrollment are excluded, this has implications for the population to which estimates can be generalized, as the design implicitly conditions on those who survive symptom free. When they are included, they may attenuate estimates of vaccine effectiveness relative to an ideally conducted trial as presumably vaccination post symptom onset is ineffective at preventing illness. In a trial, because eligibility is assessed prior to randomization, participants can be screened independent of their vaccination strategy and thus effect estimates remain unbiased. However, in an observational study this event is observed only among those who are vaccinated and therefore bias may result if they are differentially excluded.

Finally, another challenge specific to observational studies is the lack of an unambiguous assignment to a vaccination strategy at time zero [8]. In a trial, participants are explicitly assigned to either vaccine or no vaccine (or placebo) at the time of enrollment and prospectively followed. By contrast, in an observational study, a participant’s vaccination strategy is often defined retrospectively by what they do (or do not do) over the follow up period (middle panel, Figure 1). When there are delays in receiving a vaccine, this creates the possibility of bias due to *immortal time* among the vaccinated as they have survived symptom-free long enough to become vaccinated [9], whereas the unvaccinated are defined independently of their survival time.

In a trial, the challenges posed by overlap between when vaccines are received and the incubation period can be addressed through careful design and a clear protocol, for instance by specifying a window in which people are eligible to be vaccinated, screening based on uniform criteria at enrollment, and having unified time zero for all strategies. In an observational study, these fixes are often unavailable. However, these challenges can still be resolved through careful consideration of the trial that one would like to perform, but cannot, and then attempting to emulate it in the observational data (bottom panel, Figure 1).

## 3 Specifying the target trial

### 3.1 Set up and notation

We consider the emulation of a target trial designed to estimate the effect of postexposure vaccine therapy on the Δ-day risk of clinical disease. The time index *t* denotes days since exposure to a case. We have available observational data *O* = (*L*_0_, *A*_0_, *D*_1_ …, *L*_Δ*−*1_, *A*_Δ−1_, *D*_Δ_, *X*^*\**^, *T* ^*\**^) on participants, where *L*_*t*_ is a set of time-varying covariates and *L*_0_ includes all covariates prior to time zero (i.e. pre-exposure). We define the following variables:

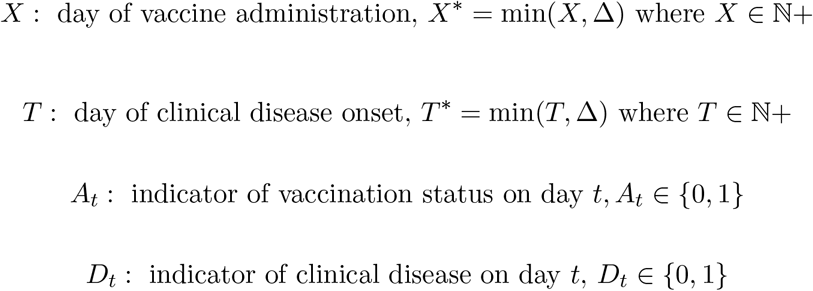

Note that under these definitions, when *X < x* then *A*_*x*_ = 1 and *T <* Δ implies *D*_Δ_ = 1. We bin both vaccination time and symptom onset time into days since exposure and administratively censor after Δ days postexposure (i.e. those unvaccinated during follow up have *X* = Δ and those without clinical disease have *T* = Δ). The trial outcome *Y* is the development of clinical disease within Δ days postexposure, i.e. *Y* = *D*_Δ_. For clarity, we make a few simplifying assumptions, although extensions that relax them are possible. First, we assume that the vaccine itself does not cause mild symptoms that can be mistaken for clinical disease. Second, we assume that the timing of the primary exposure event is measured without error and unambiguously defined. Third, we assume the goal of postexposure vaccination is the prevention of clinical disease in those exposed rather than reduction in disease severity or risk of further transmission, although in both cases the conceptualization of the target trial may be similar.

### 3.2 Possible trial designs

Under the theory that the earlier a vaccine is administered postexposure the better, the ideal causal quantity of interest, in terms of maximizing effectiveness, is likely

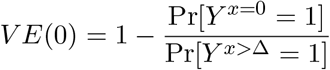

where *Y* ^*x*=0^ is a counterfactual indicator of symptoms within Δ days under *immediate* postexposure vaccination on day 0 and *Y* ^*x>*Δ^ is the counterfactual outcome under no vaccination over follow up (Note, using our definition of time-varying treatment *A*_*t*_, we could alternatively write this as 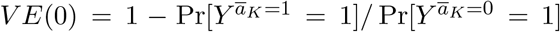 where *K* = Δ *−* 1). In a randomized controlled trial with perfect adherence, this quantity could be estimated by recruiting eligible participants immediately postexposure, randomizing them to receive vaccine or no vaccine, and comparing Δ-day incidence of symptoms in the two groups (we discuss estimating vaccine effectiveness based on the hazard ratio rather than cumulative incidence in section A.10 of the Appendix).

Alternatively, if the goal was to estimate vaccine effectiveness by day, we could imagine a design in which participants are still enrolled immediately postexposure and randomized to vaccine or no vaccine, but then further randomly assigned a day that they are to receive a vaccine. In this case our casual contrast of interest is the *t*-specific vaccine effectiveness

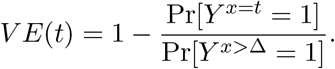

Such a design permits the estimation of the maximum delay window beyond which population effectiveness falls below a minimum threshold (see section A.11 of the Appendix). However, several challenges prevent both trials from being conducted in practice. Most importantly, the timing of enrollment and vaccination are rarely within the control of the investigator due to delays in identifying those exposed, referring them to care, and accessing a vaccine. Even if these delays were reduced in a controlled environment, that setting may not reflect how vaccines are actually administered in clinical practice and therefore less informative about real world effectiveness.

Another design, which allows participants to present “naturally”, is to specify a fixed time window that participants are eligible for enrollment and randomize them on the postexposure day they present. Given that length of delay is likely a strong determinant of effectiveness, we could improve efficiency by blocking eligible participants on the postexposure day they present. Such a design targets the *t*-specific vaccine effectiveness among those presenting symptom-free, i.e.

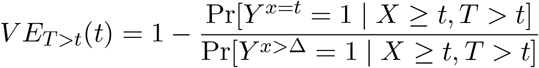

by comparing vaccine and no vaccine groups within enrollment strata. Note that, in general, the *t*-specific vaccine efficacies, *V E*_*T >t*_(*t*), will not be the same as the *V E*(*t*) defined previously as they are conditional on surviving symptom-free. Because participants are allowed to present naturally, those that present earlier may be systematically different than those presenting later with respect to their risk of developing clinical disease. Indeed, *V E*(*t*) and *V E*_*T >t*_(*t*) will only coincide when there is no effect modification by enrollment day or symptom onset time, which are both implausible. Absent this, the two measures of VE answer fundamentally different policy questions. The first, *V E*(*t*), answers: *at the time of exposure* how effective would a vaccine be if administered after a *t*-day delay? The second, *V E*_*T >t*_(*t*), answers the question: *given that an individual presents symptom-free on day t*, how effective would receiving a vaccine now be versus not?

A final design possibility is to enroll participants immediately but allow a *grace period* [10, 11], *i*.*e. a fixed time window after randomization in which vaccination can be initiated. We discuss designs that allow for a grace period further in section A*.*3 of the Appendix*.

### 3.3 Example protocol for a target trial of a postexposure Mpox vaccine

To illustrate a postexposure trial emulation, here we outline a protocol for a target trial to evaluate the effectiveness of the JYNNEOS vaccine as postexposure prophylaxis against development of symptomatic mpox infection. For simplicity, we focus on a single design: a trial with a fixed enrollment period in which participants are randomized on the postexposure day they present.

The human mpox virus (MPXV) is an orthopox virus and related to the virus that causes smallpox. In April 2022, an outbreak of mpox occurred in several countries prompting the World Health Organization to declare a public health emergency of international concern [12]. A two-dose live replicating vaccine for smallpox and mpox (MVA-BN), licensed as JYNNEOS™, was approved by the Food and Drug Administration (FDA) in 2019. During the outbreak, the vaccine was offered as postexposure prophylaxis to contacts of confirmed mpox cases. In guidance documents, the U.S. Centers for Disease Control and Prevention (CDC) recommended that unvaccinated people exposed to the mpox virus be vaccinated with a first vaccine dose against mpox within 4 days of exposure for the greatest likelihood of preventing disease [13], though also suggested there may still be benefit to vaccination within 14 days of exposure [14, 15]. Licensure of JYNNEOS was supported by animal studies [7, 16–18] and immunogenicity studies [19] but to date no trial data on the postexposure effectiveness of the vaccine against mpox exists. Therefore, an emulation of a postexposure trial using observational data may provide useful evidence for setting policy.

Table 1 gives an overview of the target trial protocol to estimate *V E*_*T >t*_. In Appendix section A.4, we provide further description of each component of the protocol.

**Table 1:** Example protocol for the specification and emulation of a target trial of postexposure vaccination for prevention of mpox.

| Protocol component | Target trial specification | Emulation |
| --- | --- | --- |
| Eligibility | High<sup>a</sup> or intermediate<sup>b</sup> risk exposure to a PCR-confirmed mpox case within the first 14 days postexposure AND negative PCR for mpox or orthopox virus at enrollment AND no symptoms AND no prior history of JYNNEOS vaccination | same |
| Treatment strategies | (1) JYNNEOS vaccination immediately upon enrollment<br>(2) no JYNNEOS vaccination during 21 days postexposure | same |
| Treatment assignment | non-blinded 1:1 random assignment to either (1) or (2) at enrollment | same but randomization is emulated by conditioning on covariates and time since exposure |
| Outcomes | 21-day cumulative incidence of disease defined as symptom onset and PCR-confirmed mpox or orthopox | same |
| Time zero | Day enrolled and assigned a strategy during the 14-day fixed enrollment period starting from exposure date | emulated as a sequence of nested trials starting each of the 14-day enrollment period |
| Follow up | Start at time zero and follow until clinical disease onset, loss to follow up, or 21 days have elapsed, whichever is first | same |
| Causal contrast | Intention to treat (ITT)<br>Per protocol | observational analog of per protocol effect |
| Statistical analysis | ITT: compare cumulative incidence of clinical disease under each strategy, adjusting for loss to follow up and prognostic factors to increase efficiency<br>Per protocol: Use IPW/g-formula/ g-estimation to account for non-adherence. | same as per protocol |

<sup>a</sup> *High risk*: direct mucosal or broken skin contact with lesions or bodily fluids OR any sexual or intimate mucosal contact OR indirect mucosal or broken skin contact with lesions or bodily fluids via linens, clothing, or other materials.
<sup>b</sup> *Intermediate risk*: unmasked exposure to respiratory droplets (within 6 ft for >3 hours) OR direct contact between intact skin and lesions or bodily fluids OR indirect contact between intact skin and lesions or bodily fluids via linens, clothing, or other materials OR indirect contact between exposed individual's clothing with linens or bodily fluids.

## 4 Emulating postexposure trials

Once the target trial is specified, we can attempt to emulate it using observational data. Emulating a postexposure vaccination trial will generally require linking high quality case and contact surveillance with clinical databases or registries recording vaccinations as well as intensive post vaccination symptom monitoring. Here, we outline how to emulate the main components of the target trial as well as common challenges, using the JYNNEOS vaccine example to help ground our discussion. Additional details on the identifiability conditions and specific data manipulation and estimation steps necessary to emulate all trial designs discussed above are available in the Appendix (sections A.5 - A.7).

### Eligibility

Ideally, eligibility criteria in the emulation should match those in the target trial. In particular, this means we cannot include restrictions based on post-baseline events (e.g. “exclude those vaccinated more than 15 days after exposure or those vaccinated after symptoms”) as these may introduce bias and would be unavailable at baseline in the target trial. Additional challenges may arise because there is no direct contact with participants at enrollment. Rather we must rely on routinely collected data which may not be fit-for-purpose. For instance, we may have to assume that those without a previous vaccination in the electronic medical records database did not receive a vaccine from a different healthcare system.

More broadly, when emulating postexposure trials, determining eligibility requires knowing who is actually at risk of infection. This means proper classification of those exposed to an index case is needed as well as an accurate history of vaccination or previous infection and screening for symptoms or PCR-positivity at enrollment. Infection history may be spotty if it mostly consists of prior recorded infections unless the pathogen is novel or invades a mostly naive population. Vaccination history may come from medical records or vaccination registries. Ideally, contacts of the index case would all be offered PCR testing upon notification of exposure and then enrolled in active symptom tracking, such as through daily phone calls or text messages, as this would prevent differential eligibility assessments of vaccinated and unvaccinated participants. However, in practice, investigators may have to assume that the lack of a positive PCR test and/or no passive symptom report constitutes no infection at time eligibility is assessed in the emulation.

### Treatment strategies

The vaccination strategies to be emulated should also match those in the target trial. As participants in observational data sets will almost always be aware of their treatment strategy, the trial emulated will typically be a pragmatic (unblinded) trial. To emulate a target trial, we identify individuals in the database who meet all of the eligibility criteria. We then assign them to the strategy or strategies that are consistent with their observed data at baseline.

To properly “assign” participants to strategies in the emulation, accurate data on the postexposure timing of vaccination is necessary. This will also allow us to censor them when they deviate from their assigned protocol. In order to identify the unvaccinated, we must inevitably assume that those without vaccinations recorded in a registry or health records truly did not receive a vaccine during follow up. This may be a problem if participants can receive care from sources not covered by study data.

Another challenge is that, to properly define regimes, the exposure date should be accurate and unambiguously defined. The accuracy of exposure information may depend on the salience of the event and the ability of index cases or their contacts to recall interactions. An unambiguous definition requires a detailed description of what constitutes possibly infectious contact informed by the underlying biology. In our mpox example, this description comes from guidance published by the CDC, but may not be as clear for other pathogens. Participants may also be exposed multiple times or over an extended duration, in which case determining which time to set as the definitive exposure date may be less clear. As a sensitivity analysis we might consider multiple alternative definitions.

### Assignment procedures

In the emulation, allocation to treatment strategies is assumed to be random conditional on a sufficient set of covariates to control confounding. For postexposure vaccination against mpox this may include time since exposure, risk level of contact with index case, calendar week, geographic region, age, sex, gender, coexisting conditions affecting immune system (e.g. HIV or STIs, obesity, cancer, immune suppressing therapies), and proxies for healthcare utilization (e.g. flu vaccination, outpatient visits, HIV-PrEP).

In practice, our ability to correctly estimate effects will depend on the conditional randomization assumption, at least approximately, holding (equivalent to assuming that there is little residual confounding). If those who access postexposure vaccines are those with higher risk exposures to mpox or with weaker immune systems (along some dimension not captured by the covariates) then we will likely underestimate the true effectiveness of the vaccine. On the other hand, if those who access postexposure vaccines are healthier and more likely to engage in healthy behaviors more broadly (again along dimensions not captured by the covariates), then we will likely overestimate the true effectiveness of the vaccine. The availability of rich covariate information on participants as well as deep subject matter knowledge about the determinants of both who gets vaccinated and the clinical course of disease are essential.

While direct verification of this assumption is not possible, several design and analytic strategies can limit or quantify the bias that would result from violations. One strategy is to identify possible negative outcome controls [20, 21], that is outcomes where confounding structure is expected to be similar but are plausibly unaffected by vaccination. For instance, routine visits for other conditions may be a proxy for unmeasured health-seeking behaviors or testing positive for syphilis may be a proxy for unmeasured high-risk sexual behavior. Another strategy is to conduct a sensitivity analysis to quantify the potential bias by evaluating change in estimated effect across a plausible range of parameter values dictating the strength of unmeasured confounding [22].

### Outcome

Outcome definitions and measurements should be as similar to those in the target trial as possible. In a postexposure vaccine trial, there is often a regular system for monitoring of symptoms over the follow up period. In an observational emulation, this data may be passively collected, leaving the opportunity for potential outcome missclassification, particularly when there is a mild form of the disease which may go unnoticed or unreported or when participants may seek care from providers not covered by study data sources. This may be less of a concern when cases are reportable or the pathogen is novel. Existing symptom monitoring systems may be in place as part of contact tracing and testing systems in which case they can be leveraged. Ideally, ascertainment of symptoms would be blind to an individual’s vaccination status. If those who are vaccinated are better surveilled or use passive systems more frequently this could introduce bias.

### Causal contrast

In theory the contrasts will be the same as in the target trial, although in some instances a corollary of the intention-to-treat effect may not be estimable from the observational data. Here, we focus on the per-protocol [23] analysis of *V E*_*T >t*_.

### Statistical analysis

Compared to the analyses in the target trial, the analyses in the emulation are complicated by two factors. First, randomization is assumed to only hold conditional on covariates. Therefore our analysis must include an appropriate method of adjustment such as outcome regression, matching, inverse-probability weighting, or a combination thereof.

Second, unlike in a trial, in an emulation the assigned strategy at baseline is not known, rather it must be inferred from the observed data. In particular, participants are not assigned to vaccine or no vaccine at time zero. To avoid immortal time bias, we need to choose a start of follow up in the emulation in a way that ensures that the distribution of time since exposure is the same in both groups [24]. In the fixed enrollment period design, this can be accomplished via emulating nested daily sequential trials: starting from exposure date, each day we identify participants who are eligible to participate (e.g. no prior vaccination or mpox infection) and assign those receiving a vaccine on that day to the vaccine strategy and those who do not receive a vaccine on that day to the no vaccine strategy (see Appendix section A.6). In this setup, unvaccinated participants will be eligible to serve as controls in multiple trials until they receive a vaccine or develop symptoms. To estimate per protocol effects, we censor participants when their data deviates from their “assigned” regime and then adjust for possible time-varying selection bias using any g-method such as inverse-probability of censoring weights. Additionally, because we are using the same participant in multiple nested trials our observations are no longer independent. Therefore, appropriate adjustment to our standard errors is necessary to account for possible correlation across observations. Adjustment can be made either by using a cluster-robust variance estimator or the bootstrap.

## 5 Simulation

To demonstrate the benefits of the target trial approach, we simulated data from hypothetical observational study under a known data generation process in which there is an overlap between vaccination timing and the timing of symptom onset (full details in section A.12 of the Appendix). Figure 2 shows an example of the overlap in the distribution of simulated vaccination times and disease onset times when *V E* = 0. We used this setup to compare explicit emulation of a target trial with a few common estimation strategies drawn from the literature. Specifically, we compare the following strategies:

**Figure 2.**
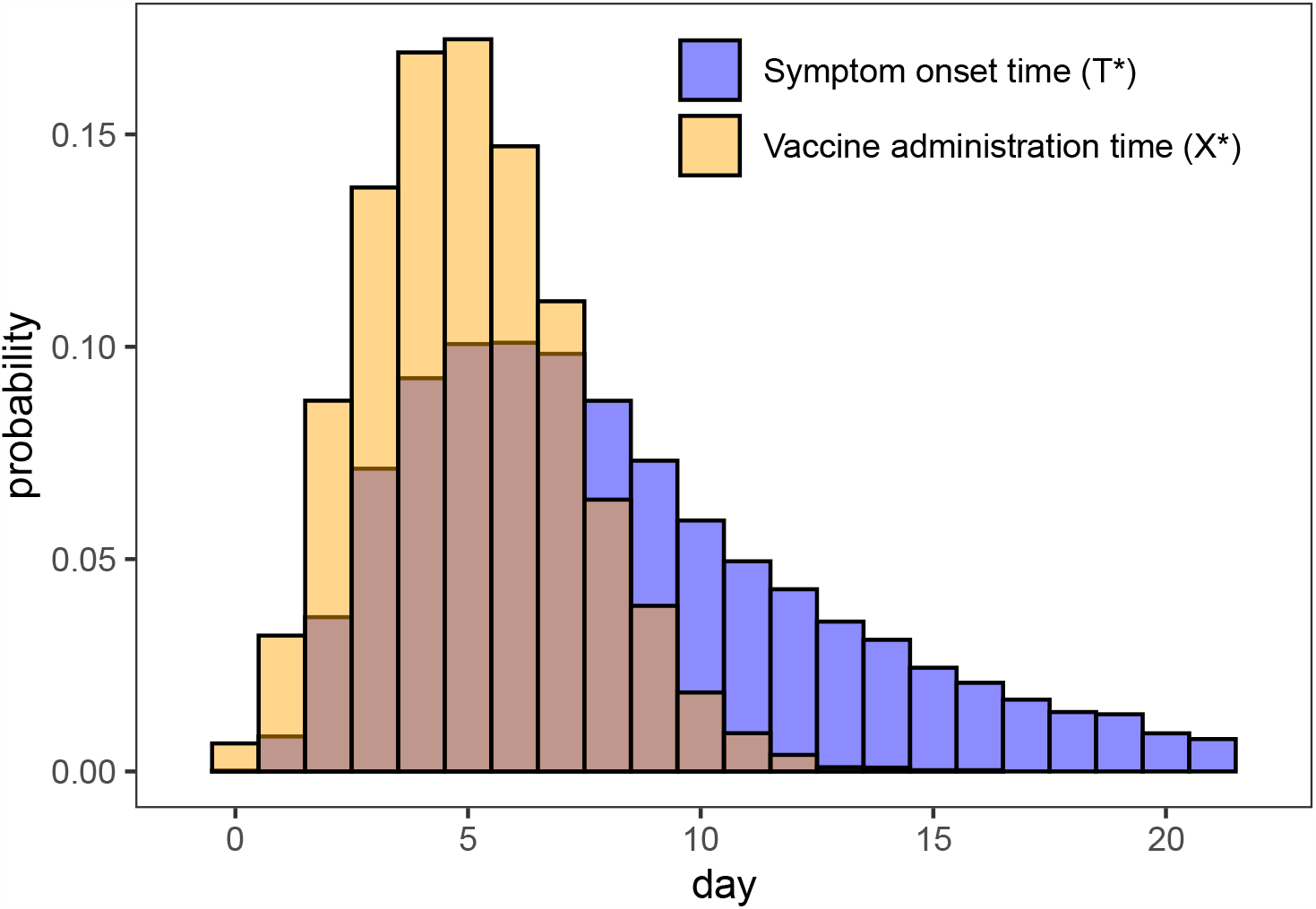
Distribution of simulated vaccination times (*X*^*\**^) among vaccinated and symptom onset times (*T* ^*\**^) among cases when *V E* = 0 over the 21 days of follow up showing the degree of overlap.

1. *naive, leave* - a simple comparison of the “ever vaccinated” and “never vaccinated” using the relative risk regression model Pr[*Y* = 1 | *X*] = exp*{β*_0_ + *β*_1_*I*(*X <* 21)*}* with vaccine effectiveness estimated as 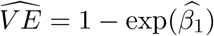.
2. *naive, move* - re-classify those who receive vaccine after developing symptoms as “unvaccinated”, i.e. we use the relative risk regression model Pr[*Y* = 1 | *X*] = exp{*β*_0_ + *β*_1_*I*(*X < T*)} where *I*(*X < T*) implies only those who receive vaccine prior to symptom onset are “vaccinated” with vaccine effectiveness again estimated as 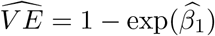.
3. *target trial* - we emulate a sequence of nested daily trials as described above and in section A.6 of the Appendix. In each trial, we censor participants when they deviate from their assigned strategy at baseline and use inverse-probability of censoring weights to adjust for selection bias. These nested trials are combined, and vaccine effectiveness is estimated using standardized cumulative incidence curves from a pooled logistic regression.

In Figure 3 we compare estimates of *V E* to the truth across two scenarios: the first when the true *V E* = 0% and the second when the true *V E* = 31.6%. Under the null, the naive approaches are upwardly biased due to immortal time bias (i.e. by definition vaccinated have to survive long enough to be vaccinated while unvaccinated are at risk at all time points), while the target trial approaches yield valid estimates. This persists in scenario 2 where *V E* = 31.6%, although the relative bias of the first approach is somewhat offset by the fact that those vaccinated after developing symptoms are included with vaccinated. In the appendix, we also compare estimation strategies when *V E* varies with timing of vaccination (Figure A7), when effectiveness is defined as one minus the hazard ratio and/or a time-varying cox model is used (Figure A8), and when the degree of overlap between vaccination and symptom onset is varied (Figure A9). Of note, in this simple scenario both the target trial emulation approach and the time-varying cox model yielded unbiased estimates of VE when defined as one minus the hazard ratio.

**Figure 3.**
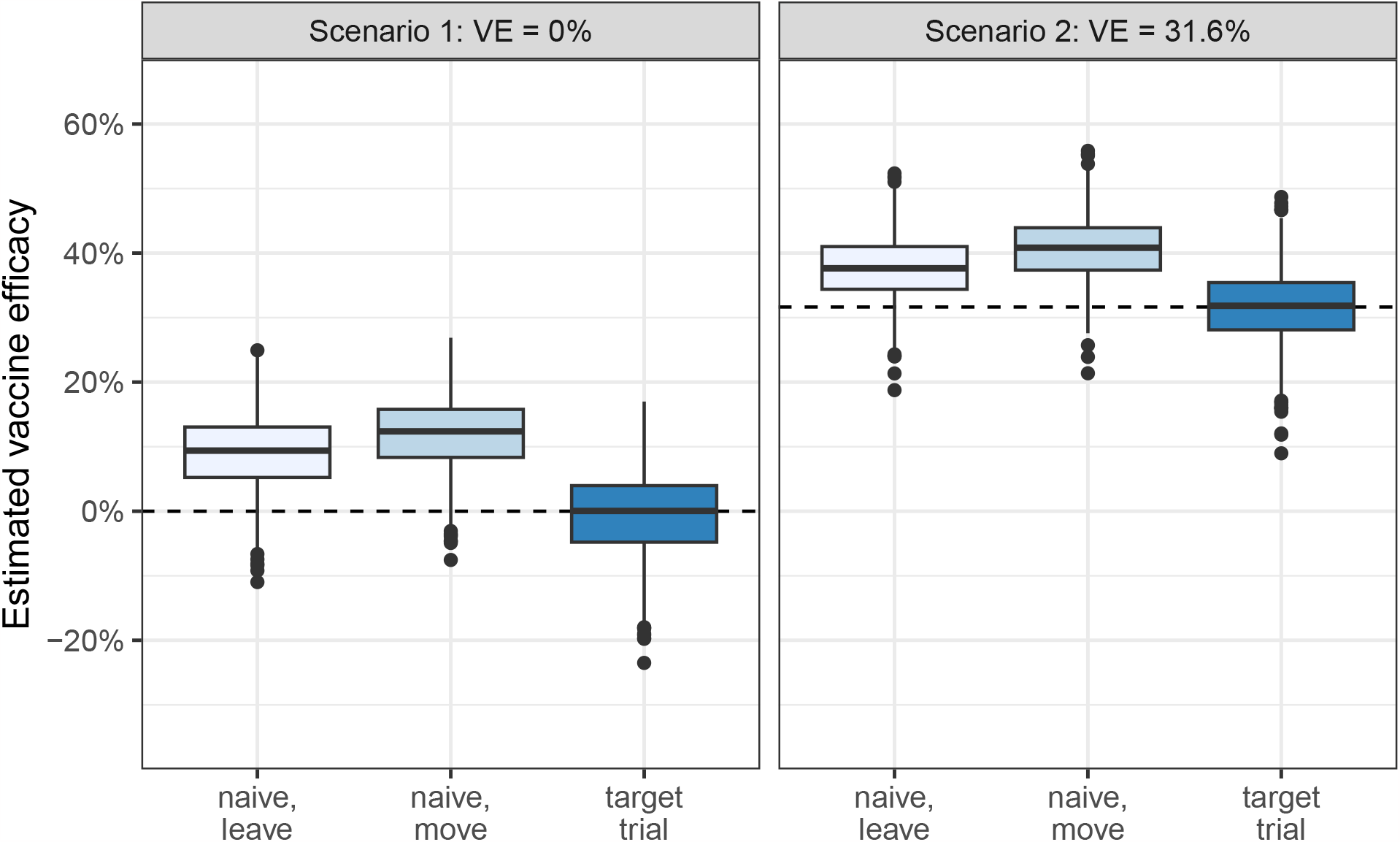
Simulated *V E* estimates compared to the truth for the three estimation strategies described in section 5. Naive, leave refers to simple comparison of those ever vaccinated over follow up versus never vaccinated. Naive, move is also a comparison of ever versus never vaccinated but those who receive vaccine after symptoms start are re-classified as never vaccinated. Target trial refers to the approach of emulating a sequence of nested trials as discussed in the main text. Based on 1000 monte carlo simulations. Dashed line shows true value in each scenario.

## 6 Discussion

Accurate assessments of postexposure effectiveness of vaccines could be useful for curbing the worst sequelae of many pathogens, but trials are often infeasible or unethical. Here, we specified target trials for postexposure vaccination and describe how to emulate them using observational data. Using the example of mpox vaccines, we discussed some of the unique challenges of emulating postexposure vaccination trials, including the central role played by the distribution of vaccination times and the incubation period. Throughout we emphasize the clarifying role of the target trial framework and conclude with simulations showing how emulating the trial can help avoid several common biases in observational analyses.

Previous studies have emulated trials of pre-exposure vaccines, particularly during the COVID-19 pandemic [25–28]. These studies filled gaps in the literature by emulating trials which were not feasible to implement in practice such as head-to-head comparisons of vaccines [26], effectiveness against new variants [27], effectiveness of boosters [28, 29], and effectiveness in important subgroups such as children [27] and the immunocompromised. Observational emulations of post-exposure vaccines could perform a similar function.

We considered postexposure trials where the goal of vaccination is to prevent the onset of clinical disease. However, other goals such as reducing severity or transmission are also possible. In rich observational datasets multiple primary and secondary endpoints may be feasible. To emulate trials in which the goal is to reduce severity, one could simply replace onset with an alternative outcome such as hospitalization or death due to disease of interest in the trials outlined above. Emulating a trial of the effect of postexposure vaccination on transmission would require close follow up or even random testing of the contacts of the vaccinated and unvaccinated participants and may be compromised by changes in exposure behaviors due to lack of blinding in most observational settings. However, if PCR tests were administered to everyone independent of symptoms, effectiveness against infection (PCR-positivity) is a lower bound on effectiveness against transmission [30].

## Data Availability

Code and data are available at: https://github.com/boyercb/pep-target-trials

https://github.com/boyercb/pep-target-trials

## Web Appendix

## Appendix

### A.1 Day zero randomization designs

In the main text, we discussed two trial designs starting on postexposure day zero. In the first, participants are enrolled on postexposure day zero, randomized, and immediately receive either vaccine or no vaccine with the goal of estimating the Δ-day vaccine effectiveness in the ideal case in which there is no delay between exposure and vaccination. Under perfect adherence this trial targets the estimand

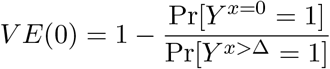

or, alternatively,

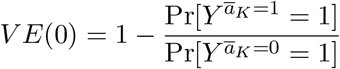

using our definition of time-varying treatment *A*_*t*_ with *K* = Δ *−* 1 and overbars representing past history, i.e. 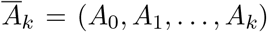. This value is likely an upper bound on vaccine effectiveness under more plausible scenarios of delay.

In the second design, participants are still enrolled and randomized on postexposure day zero, but they are then further randomly assigned a postexposure date to receive the vaccine. Under perfect adherence, the casual contrast of interest is now the *t*-specific vaccine effectiveness

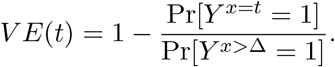

or, alternatively,

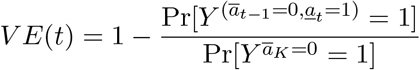

with underbars representing future history, i.e. *<u>A</u>*_*k*_ = (*A*_*k*_, *A*_*k*+1_, …, *A*_Δ*−*1_). This could be used, for instance, to determine the time window public health officials and policymakers should advise individuals at risk of exposure to seek vaccination within if they are exposed (see Section A.11).

### A.2 Fixed enrollment period designs

Also mentioned in the main text, when the timing of vaccination is not under the strict control of the investigator, a possible design is to specify a fixed time window in which participants are eligible to be vaccinated and randomize them on the postexposure day they present. Under perfect adherence, this design could then target the *t*-specific vaccine effectiveness among those presenting symptom-free, i.e.

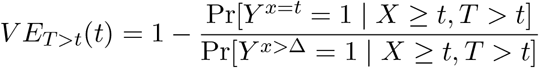

or

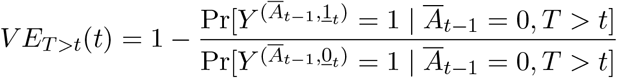

by comparing vaccine and no vaccine groups within enrollment strata. Note that, in general, the *t*-specific vaccine efficacies, *V E*_*T >t*_(*t*), targeted in this trial will not be the same as the *V E*(*t*) defined previously as they are conditional on presentation time and being symptom-free at enrollment. More often, in practice, the *t*-specific estimates *V E*_*T >t*_(*t*) are pooled together into a weighted average effectiveness over the enrollment period. However, we stress caution in interpreting pooled estimates. Because participants are allowed to present naturally rather than being assigned a time at day zero, those that present earlier may be systematically different than those presenting later with respect to their risk of developing clinical disease. Therefore the pooled estimates are among a subpopulation who survive symptom-free and may not generalize to other populations with different propensities for delay.

### A.3 Adding a grace period

An alternative to the day zero design which also allows for delays in vaccination but doesn’t require consideration of all possible delay regimes is to specify a *grace period*, i.e. a fixed time window after randomization in which vaccination can be initiated. For example, in a postexposure trial of a varicella vaccine, investigators might randomize sibling contacts of an index case to vaccine or placebo and then specify contacts are adherent if they receive their assigned treatment within *within 72 hours* of the appearance of the first skin lesion in the index case. Under this design, the causal target would be the average vaccine effectiveness if received during the *m* days of the grace period, i.e.

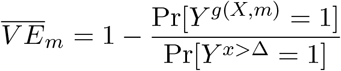

where

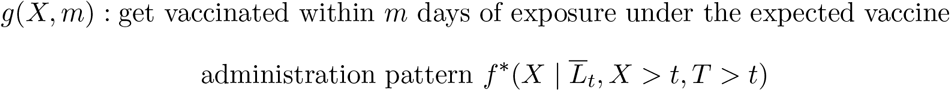

and where, for instance, in the hypothetical varicella trial *m* = 3. Although in theory randomization could occur on any postexposure day followed by *m*-day grace period, in practice grace periods starting from randomization on day zero probably make the most sense. When effectiveness varies by the time since exposure, as it most certainly does for postexposure vaccines, a grace period design estimates the average effectiveness *under the “natural” course of vaccination over the grace period*, i.e.

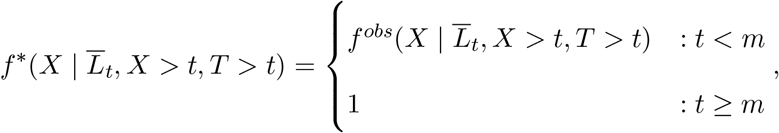

or alternatively

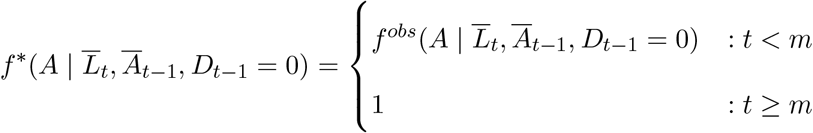

using time-varying treatment *A*_*t*_ instead of *X* and time-varying disease indicator *D*_*k*_ in place of *T*. We note the effectiveness under other stochastic strategies may be possible under additional assumptions [1]. This implies that two trials identical in all respects except for the distribution of vaccinations over the grace period could yield substantially different estimates. Therefore, a trialist pursuing this design has to strike a balance when defining a grace period between ensuring the period is short enough that benefit is immunologically possible and the trial is adequately powered, but also long enough that the regime is clinically feasible under reasonable assumptions about how quickly patients are notified of their exposure to a case and can access a vaccine in the real world. If properly conceived, a grace period design can provide evidence about average effectiveness of postexposure vaccination administered within a certain window under real world conditions. As such it may be a more useful estimate for population planning or modeling studies than those produced by the fixed enrollment period design above.

### A.4 Example target trial specification for JYNNEOS vaccine

#### Eligibility

Individuals over 18 years of age who had an intermediate or high risk exposure to a person with laboratory confirmed mpox case, no history of JYNNEOS vaccination, no positive PCR for mpox or other orthopox virus at enrollment, and who were referred within 14 days of exposure are eligible for this study. We use the CDC definitions of high and intermediate risk exposures [2] for mpox (Table 1).

#### Treatment strategies

1. a single JYNNEOS vaccination dose (either the intradermal or subcutaneous regimen) at enrollment and 2) no mpox vaccination dose over the 21-day follow up period.

#### Assignment procedures

Individuals are randomly assigned to one strategy within permuted assignment blocks defined by day of presentation at the clinic and possibly other covariates of interest. Individuals are aware of the strategy to which they have been assigned (unblinded).

#### Outcomes

The primary outcome is PCR-confirmed mpox or orthopox infection within 21 days of exposure. Secondary outcomes could include disease severity or safety endpoints.

#### Follow-up period

Follow-up begins at date of exposure to the index case and ends at either the occurrence of the outcome, 21 days after exposure, or loss to follow-up, whichever occurs first (We discuss the possibility of delayed outcomes based on biology and their implications for observational emulations in Appendix section A.9).

#### Causal contrasts

Intent-to-treat and per protocol effects of JYNNEOS vaccination.

#### Statistical analysis

In the intention-to-treat (ITT) analysis, we compare the cumulative incidences in each group defined by assignment and calculate the vaccine effectiveness as 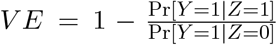 where *Z* is an indicator of random assignment to strategy (1) or (2). In the stratified design, we can either calculate ITT effects for the *t*-specific vaccine efficacies separately or, under additional assumptions, pool together into a 14-day average. Cumulative incidence curves can be estimated in each arm via the Kaplan-Meier estimator or a pooled logistic model. Loss to follow-up can bias estimates of ITT effects [3]. We can adjust for the resulting selection bias under the assumption that measured covariates (in practice often just baseline) include all determinants of loss to follow-up and the outcome.

The per-protocol analysis is similar to the ITT analysis except that individuals are censored if they deviate from their assigned treatment strategy, e.g. by declining the vaccine when assigned to vaccine or obtaining it outside of the trial if assigned to no vaccine. We can adjust for the possible time-varying selection bias due to censoring for protocol deviations (and/or loss to follow-up) through an appropriate g-method under the assumption that the measured variables include approximately all determinants of adherence and the outcome. 95% confidence intervals may be estimated via bootstrapping.

### A.5 Identifiability conditions

Here, we review the conditions under which data from an observational emulation can be used to identify the causal contrasts of interest in the target trial designs discussed above. As previously, let 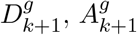, and 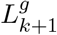 represent the counterfactual outcome, vaccination, and covariate history under vaccination strategy *g*. To identify effects, the following conditions must hold for *k* = 0, *…*, Δ *−* 1:

1. *Consistency:* If 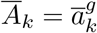 then 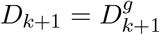 and 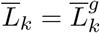.
2. *Sequential exchangeability:* 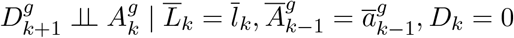
3. *Positivity:* 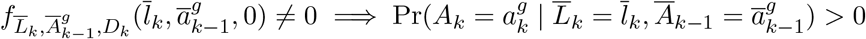

The first condition requires that our vaccination strategies are sufficiently well-defined such that they match an intervention that would be assigned in the target postexposure trial and are reflected in the observed vaccination patterns in our observational study. The second condition is alternatively known as the “no unmeasured confounding assumption” and requires that sufficient covariate information is collected in the observational study such that potential outcomes are as if randomized, i.e. conditionally independent, given past vaccination and covariate history. The final assumption requires that there is a positive probability of receiving a vaccine in each covariate strata.

In Figure A1, we use Single World Intervention Graphs (SWIGs) to represent the postexposure process, i.e. time-varying evolution of vaccination, symptoms, and covariates. Because SWIGs explicitly depict potential outcomes under interventions they are well-suited to reasoning about exchangeability conditions. On a SWIG, a potential outcome 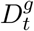 is conditionally independent of treatment *A*_*t*_ if there are no unblocked *backdoor* paths connecting 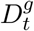 and *A*_*t*_ conditional on past treatment and covariate history. Using SWIGs helps us illuminate a subtle point regarding the conditional exchangeability assumption under the grace period strategy compared to the static strategies in the fixed enrollment period and day zero designs, which is developed further in [1]. Specifically, when there is an unmeasured common cause of vaccination *U*_*A*_ at any two time-points exchangeability will hold for the static regimes of the fixed-enrollment and day zero designs, but it will not hold for the grace period under natural initiation of treatment.

**Figure A1:**
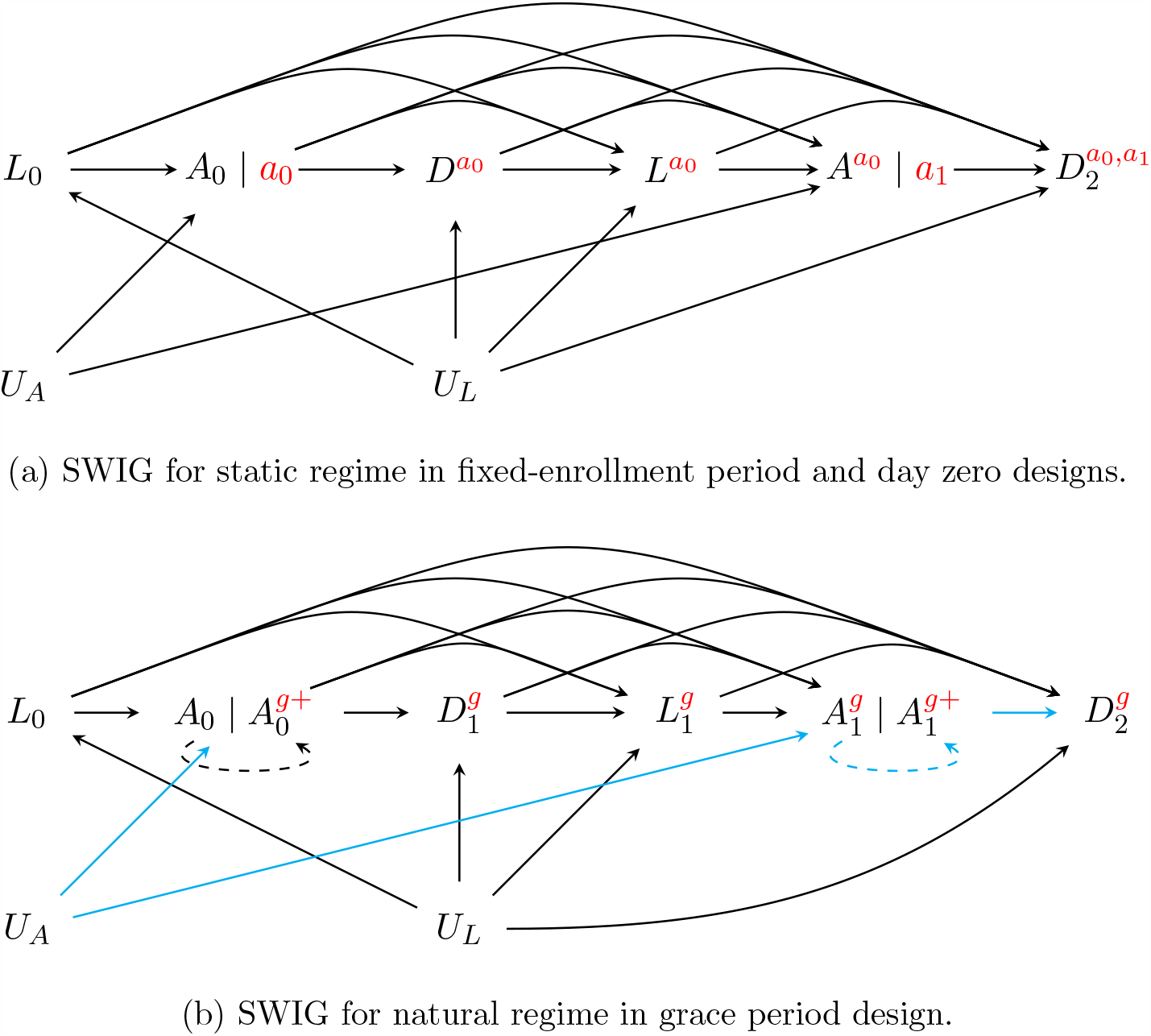
Causal graphs depicting the postexposure process under different interventions. The variables *L*_0_, *L*_1_, *A*_0_, *A*_1_, *D*_1_, and *D*_2_ are as defined in the main text. Unmeasured variable *U*_*L*_ is a common cause of measured covariates and the outcome (symptoms) and unmeasured variable *U*_*A*_ is a common cause of vaccination. The blue line highlights the open backdoor path which leads to a failure of the sequential exchangeability assumption.

### A.6 Data setup and additional emulation details

Here, we demonstrate the data manipulation steps required to emulate the three trial designs —day zero, fixed enrollment period, and grace period— discussed above using observational data. Our goal is to emulate the analysis that would have been conducted in the ideal trial assuming that the necessary identifiability conditions are valid in the observational study.

Two crucial differences between a randomized trial and an observational study are that 1) the former has a well defined start of follow up, or *time zero*, from which study outcomes are assessed and 2) all participants are assigned a particular treatment strategy. By contrast observational studies generally do not have a uniquely defined time zero and participants may have data consistent with multiple treatment strategies. In each design, certain data manipulation steps are applied to the observational data to solve these issues.

#### A.6.1 Fixed enrollment period trial emulation

When emulating a fixed enrollment period design, the issue is that participants in the observational data generally meet the eligibility criteria at multiple time points, and therefore there is no unambiguous time zero from which to start follow up. For instance, consider a postexposure vaccination trial in which participants are eligible anytime in the first 5 days after exposure if they have no previous vaccination history and no symptoms at presentation. In a real trial the participant would be enrolled and randomized on a particular day and that would be their time zero. In the observational data, a participant may meet these criteria continuously, for instance between days 0 and 4. The question is then when should their follow-up start? On day 0, 1, 2, 3, or 4? The choice has to be applied equivalently to vaccinated and unvaccinated participants to avoid *immortal time bias*. One possibility is to randomly choose a start time among the days they are eligible. However, a more efficient choice is to use every eligible time by emulating a sequence of multiple nested target trials each with a different start time. A natural choice for postexposure vaccination for a pathogen with a relatively short incubation period is to emulate a series of daily nested trials, i.e. on day zero condition on those who meet the eligibility criteria and compare those who are vaccinated on that day to those who are unvaccinated on that day, and then repeat on all days within the fixed enrollment period (schematic Figure A2). Participants in the observational study can be enrolled in trials starting on multiple days as long as they meet the eligibility criteria at time zero. As suggested in [4], this design is akin to estimating the parameters of the structural nested model

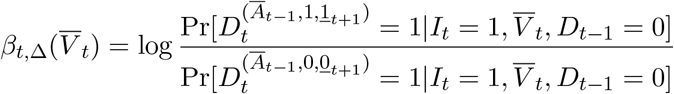

where *I*_*t*_ is an indicator that a participant is eligible for the trial starting on day *t* and *V*_*t*_ are a subset of covariates which may be effect modifiers.

To demonstrate the required data manipulation steps, consider the six individuals shown in Table A1 with vaccination and symptom onset times recorded during a hypothetical observational study. To emulate a trial with a fixed five day enrollment period postexposure, we create one copy of the dataset for each trial day. Then in each copy we apply the proper eligibility criteria (e.g. individuals should be disease-free and not vaccinated on a previous day) and assign those vaccinated on that trial day to be “vaccinated” and those who have not been vaccinated yet to be “unvaccinated”. For example, individual 2 in Table A1 is vaccinated on on day 2 and doesn’t develop symptoms, therefore in the emulation they will participate in 3 trials (i.e. those starting on postexposure day 0, 1, and 2). In trials starting after postexposure day 2 they are no longer eligible because they have already been vaccinated. In each trial, follow up time is adjusted to start on the postexposure day of interest and end either at symptom onset or at the maximum follow up day which may be fixed from the index exposure day or be of fixed length from the trial day. In intention-to-treat analyses, participants are “assigned” based on their baseline status in the nested trial and followed throughout regardless of whether they later deviate. In per protocol analyses, individuals in each nested trial are censored when they deviate from their baseline assignment in that trial. For example, individual 3 in Table A1 is unvaccinated in trials starting on days 0 through 3, but in each of these trials is censored on day 4 in the per protocol analysis because they deviate from their baseline assignment by becoming vaccinated.

#### A.6.2 Day zero trial emulation

When emulating a day zero trial in which participants are randomized to a particular delay before receiving a vaccine after exposure, the issue is that participants in the observational data will have data consistent with multiple delay regimes. Consider a trial where participants are randomized on day zero to one of the following strategies: (1) receive vaccine on day zero, (2) receive vaccine on day one, (3) receive vaccine on day two, (4) receive vaccine on day three, or (5) to receive no vaccine over the follow up period (schematic Figure A3). In a real trial participants would be assigned to one of the five regimes at the start. In the observational data, however, some individuals will get vaccinated on day 0 and therefore only have data compatible with the first strategy, but others will not get vaccinated on day zero and will have data compatible with multiple strategies at baseline. The question is now which strategy should we assign them to? As with the start time in the previous design, one option is to pick a single strategy at random from the strategies their data is consistent with. However, the more efficient choice is to assign them to all possible strategies by creating exact copies —often called clones— of any individual whose data is consistent with multiple regimes and assign each of their clones to a different strategy.

**Table A1:** Enrollment of six hypothetical individuals in daily nested trials for 5-day vaccination window based on observed data.

| ID | Vaccination<br>(1: yes, 0: no) | Day of<br>vaccination | Clinical disease<br>(1: yes, 0: no) | Day of<br>disease onset | No. of trials<br>enrolled |
| --- | --- | --- | --- | --- | --- |
| 1 | 1 | 0 | 0 | - | 1 (day 0) |
| 2 | 1 | 2 | 0 | - | 3 (days 0-2) |
| 3 | 1 | 4 | 1 | 8 | 5 (days 0-4) |
| 4 | 1 | 7 | 1 | 2 | 2 (days 0-1) |
| 5 | 0 | - | 0 | - | 6 (days 0-5) |
| 6 | 0 | - | 1 | 5 | 5 (days 0-4) |

To demonstrate the required data manipulation steps, let’s return to the six hypothetical individuals from Table A1, but now in Table A2 we will use their data to emulate a day zero trial in which participants are randomized to strategies (1)-(5) in previous paragraph. Starting with the first individual, they are vaccinated on day zero and therefore have data consistent only with strategy (1), thus they are not cloned. The second individual, however, is not vaccinated until day 2 and therefore at time zero they have data consistent with any of the strategies (1)-(5), thus we make five clones of the second individual by copying their data five times and assigning each observation to a different regime. We then follow each clone forward and censor them when they deviate from their assigned regime. For instance, we know the second individual is vaccinated on day 2, therefore on day 0 we censor the clone assigned to strategy (1) because they were not vaccinated on that day. Likewise, on day 1 we censor the clone assigned to strategy (2) because they were not vaccinated on that day either, then on day 2 we censor all the remaining clones except the one assigned to strategy (3). Importantly, if the individual has symptoms before a clone is censored, as is the case for the strategy (3) and (4) clones for individual 4, then all clones will have symptoms and therefore the case is assigned to all strategies. This multiple allocation of events prevents the bias that could arise if events occurring during the delay period are systematically assigned to one of the five strategies only.

**Table A2:** Emulation of day zero trial with five vaccination delay strategies using data from six hypothetical individuals.

| ID | Vaccination<br>(1: yes, 0: no) | Day of<br>vaccination | Clinical disease<br>(1: yes, 0: no) | Day of<br>disease onset | No. of clones<br>(regimes) |
| --- | --- | --- | --- | --- | --- |
| 1 | 1 | 0 | 0 | - | 1 (1) |
| 2 | 1 | 2 | 0 | - | 5 (1-5) |
| 3 | 1 | 4 | 1 | 8 | 5 (1-5) |
| 4 | 1 | 7 | 1 | 2 | 5 (1-5) |
| 5 | 0 | - | 0 | - | 5 (1-5) |
| 6 | 0 | - | 1 | 5 | 5 (1-5) |

#### A.6.3 Grace period trial emulation

Finally, when emulating the grace period design the issues are similar to those in the day zero trial in which participants are randomized to a delay strategy, i.e. some participants in the observational study have data consistent with multiple regimes. Consider a trial where participants are randomized at day zero to either (1) receive vaccine sometime within the first five days postexposure or (2) to receive no vaccine over the follow up period (schematic Figure A4). As before, if the trial were actually conducted everyone would have an unambiguous assignment at time zero. However, in the observational data individuals who receive vaccine after day zero have data consistent with both regimes in the period before receiving the vaccine. This is important when considering some individuals may acquire symptoms prior to receiving vaccination during the grace period, in which case one might ask oneself to which strategy should they be assigned? The solution, as before, is to create clones when individuals have data consistent with multiple regimes, assign each clone to a regime, and then censor them if they deviate from their assigned regime.

To demonstrate the required data manipulation steps, we return again to the same six hypothetical individuals, but now in Table A3 we will use their data to emulate a trial with a five day grace period (i.e. *m* = 5). Starting with the first individual, they are vaccinated on day zero and therefore have data consistent only with strategy (1) and therefore they are not cloned. The second individual, however, is not vaccinated until day 2 and therefore at time zero has data consistent both strategies (1) and (2), thus we make two clones of the second individual by copying their data and assigning each observation to one of the two regimes. We then follow each clone forward and censor them when they deviate from their assigned regime. For instance, we know the second individual is vaccinated on day 2, therefore on day 2 we censor the clone assigned to regime (2), i.e. receive no vaccine over the follow up period. Again, if the individual has symptoms before any clone is censored, as is the case for individual 4, then all clones will have symptoms and therefore the case is assigned to all strategies strategies. This double allocation of events prevents the bias that could arise if events occurring during the grace period are systematically assigned to one of the two strategies only.

**Table A3:** Emulation of trial with five day grace period vaccination using data from six hypothetical individuals.

| ID | Vaccination<br>(1: yes, 0: no) | Day of<br>vaccination | Clinical disease<br>(1: yes, 0: no) | Day of<br>disease onset | No. of clones<br>(regimes) |
| --- | --- | --- | --- | --- | --- |
| 1 | 1 | 0 | 0 | - | 1 (1) |
| 2 | 1 | 2 | 0 | - | 2 (1-2) |
| 3 | 1 | 4 | 1 | 8 | 2 (1-2) |
| 4 | 1 | 7 | 1 | 2 | 2 (1-2) |
| 5 | 0 | - | 0 | - | 2 (1-2) |
| 6 | 0 | - | 1 | 5 | 2 (1-2) |

**Figure A2:**
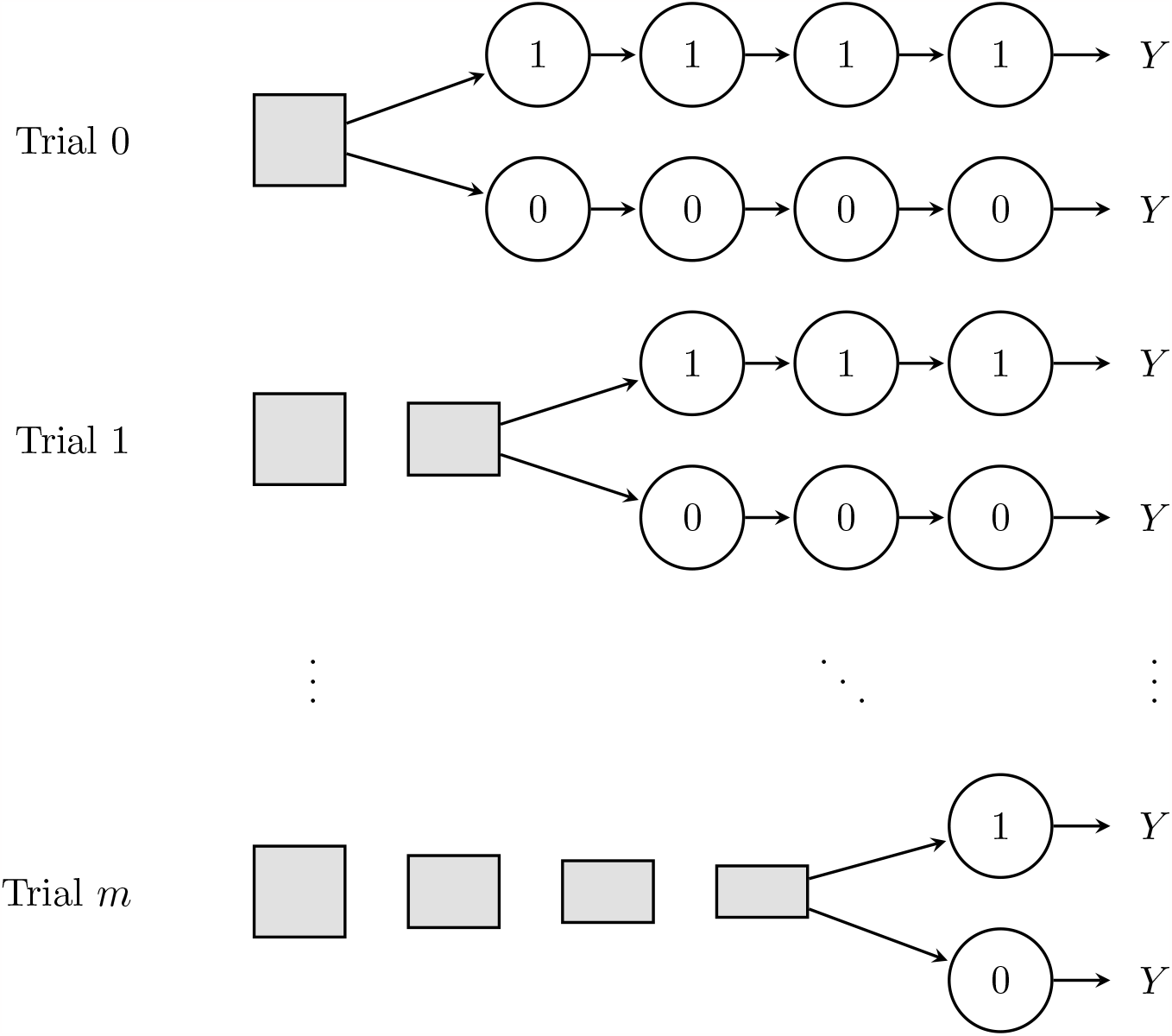
Schematic of nested daily trial design, on each day participants are assigned to vaccine or no vaccine, conditional on their history up to that day. Squares represent a pool of participants eligible for randomization and get smaller to show that the size of the pool shrinks as the number of eligible participants decreases due to symptom onset and previous vaccination; circles represent the status of individuals who have been randomized. Time moves from left to right.

**Figure A3:**
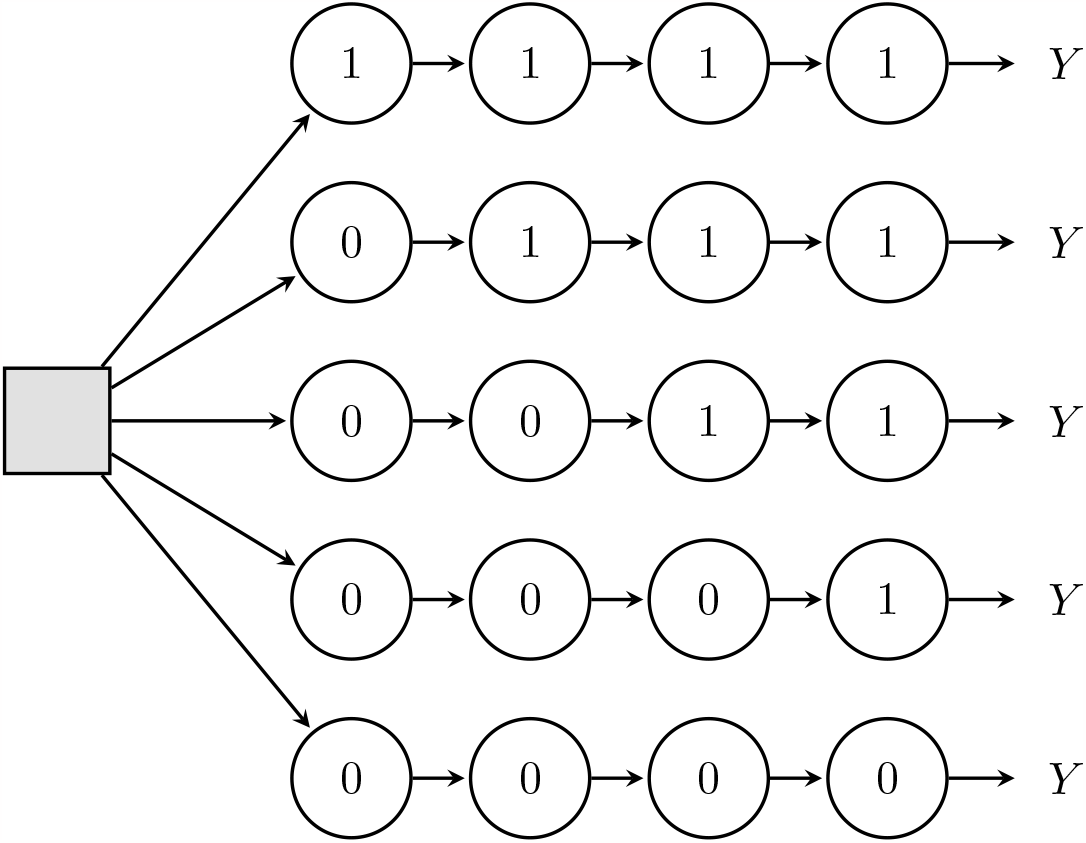
Schematic of a trial in which participants are randomized on day zero to a specific vaccination delay strategy. In the figure, on day zero, participants are assigned to either (1) receive vaccine on day 0, (2) receive vaccine on day 1, (3) receive vaccine on day 2, (4) receive vaccine on day 3, or (5) do not receive vaccine during the follow up period, with assignment probabilities allowed to vary between strategies. This could also be done by first randomizing participants to vaccine or no vaccine and then randomizing the day of vaccination among those assigned vaccine.

**Figure A4:**
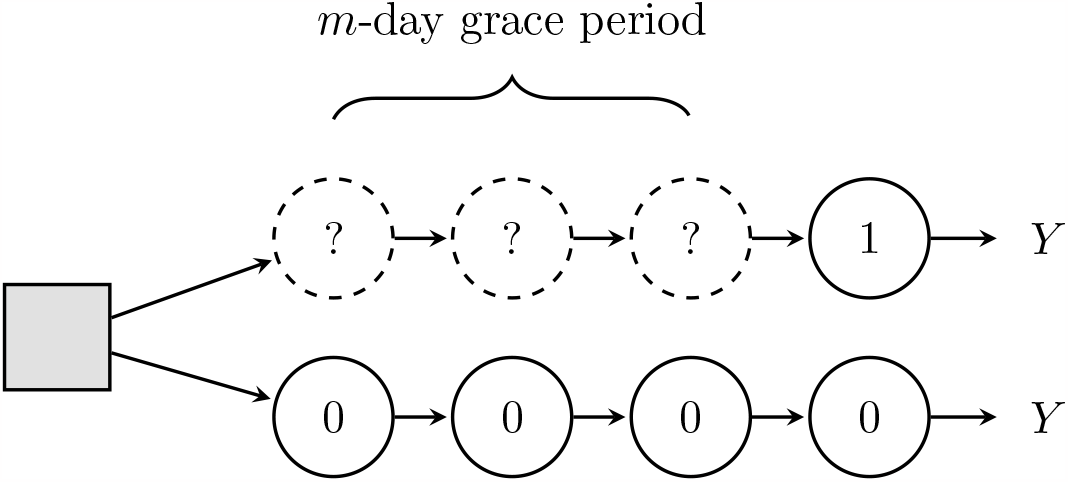
Schematic of trials with a grace period, participants are randomized to a strategy starting on day zero and given a fixed length time window in which they can initiate and then sustain thereafter.

### A.7 Estimation using inverse probability of censoring weights

Once we have completed the necessary data manipulation steps to emulate the designs above, analysis of the per-protocol effects of postexposure vaccination can be conducted. Here, we describe one possible estimation approach based on inverse probability weighting, although others are possible. The algorithm proceeds as follows:

1. For each regime of interest *Z*, define censoring indicator *C*_*k*_ which is one when the individual deviates from the assigned regime and zero when they are adherent. For example, consider a 4-day delay regime for the day zero design: if an individual is vaccinated prior to day 4, then *C*_*k*_ = 1 on the day of vaccination and *C*_*k*_ = 0 before; if they are vaccinated after day 4 or never vaccinated, then *C*_*k*_ = 1 on day 4 and *C*_*k*_ = 0 before; and if they are vaccinated as indicate on day 4, then *C*_*k*_ = 0 throughout.
2. Using the entire dataset, fit a pooled (over time) parametric regression model for the conditional probability of being censored 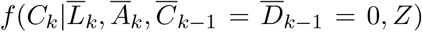. For example we could assume a pooled logistic regression model.
3. For each individual, *i*, and at each time point, *k*, in 1, …, Δ *−* 1
  a. Obtain predicted values 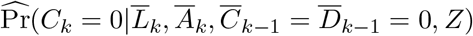.
  b. Form the appropriate weights for remaining uncensored:
    i. For the fixed-enrollment period design,

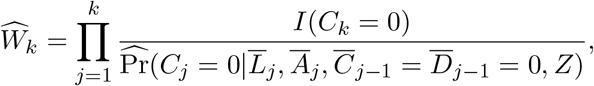

with the weights equal to 1 for those assigned to the “vaccinate” strategy (as those who initiate are always adherent).
    ii. For the day zero designs,

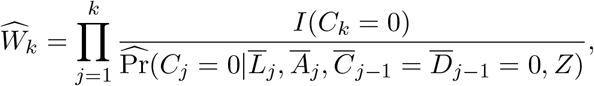

where prior to the day indicated the weights are the probability of remaining unvac-cinated and the weights after are for the probability of being vaccinated the specified day.
    iii. For a natural grace period design,

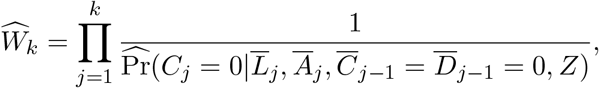

for those assigned to the “never vaccinate” strategy and

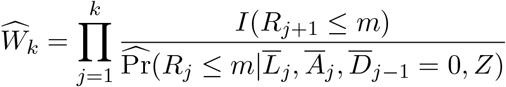

for the grace period strategy where *R*_*k*_ is the number of consecutive days prior to *k* that an individual failed to be vaccinated and

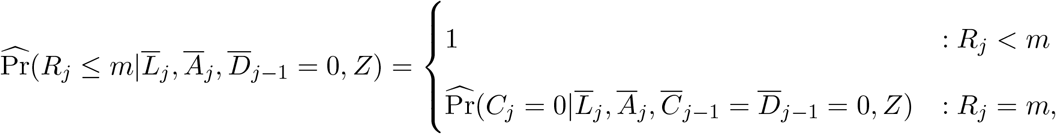
4. Using the weights 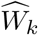, fit a suitable weighted pooled (over time) parametric regression model for the discrete-time hazard of symptom onset Pr[*D*_*k*+1_ = 1|*D*_*k*_ = 0, *L*_*k*_, *Z*]. For example, for a day zero trial, we could fit the pooled logistic regression model

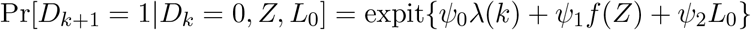

where *λ*(*k*) is the unvaccinated odds of symptom onset and *f* (*Z*) is a function of the vaccination delay. To relax assumptions on functional form, we can allow *f* (*Z*) and *λ*(*k*) to be members of a class of flexible such as restricted cubic splines.
5. Compute *V E* curve for Δ-day cumulative incidence by standardizing

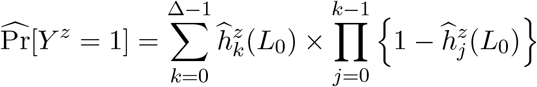

for each regime *z ∈Ƶ* where 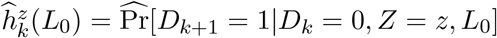 and then for any two regimes calculate 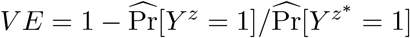.
6. For inference, draw bootstrap replicates 1, …, *B* by sampling with replacement from the dataset and repeat steps 2 through 5 to get estimates 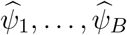. Calculate bootstrapped standard error from 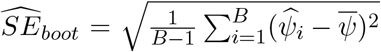. An *α*-level confidence interval may be formed via 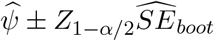.

### A.8 Hypothetical data example for estimation using inverse probability of censoring weights

**Table A4:** Data four five hypothetical individuals whose vaccination histories are consistent with at least one of: the never vaccinate strategy, the 2-day delay strategy, and a strategy to vaccinate under a grace period with *m* = 3. We define 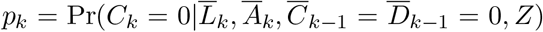 as the observed probability of remaining uncensored at time *k*.

| Person<br>($i$) | Time<br>($k$) | Vaccination<br>($A_k$) | Consec. days<br>unvaccinated<br>($R_k$) | Onset of<br>symptoms<br>($D_k$) | $W_k$ under<br>never<br>vaccinate | $W_k$ under<br>2-day<br>delay | $W_k$ under<br>grace<br>period |
| --- | --- | --- | --- | --- | --- | --- | --- |
| 1 | 0 | 0 | 0 | 0 | $\frac{1}{p_0}$ | $\frac{1}{p_0}$ | 1 |
| 1 | 1 | 0 | 1 | 0 | $\frac{1}{p_0 p_1}$ | $\frac{1}{p_0 p_1}$ | 1 |
| 1 | 2 | 0 | 2 | 0 | $\frac{1}{p_0 p_1 p_2}$ | 0 | 1 |
| 1 | 3 | 0 | 3 | 0 | $\frac{1}{p_0 p_1 p_2 p_3}$ | 0 | 0 |
| 2 | 0 | 1 | 0 | 0 | 0 | 0 | 1 |
| 2 | 1 | 1 | 1 | 0 | 0 | 0 | 1 |
| 2 | 2 | 1 | 2 | 0 | 0 | 0 | 1 |
| 2 | 3 | 1 | 0 | 0 | 0 | 0 | 1 |
| 3 | 0 | 0 | 0 | 0 | $\frac{1}{p_0}$ | $\frac{1}{p_0}$ | 1 |
| 3 | 1 | 0 | 1 | 0 | $\frac{1}{p_0 p_1}$ | $\frac{1}{p_0 p_1}$ | 1 |
| 3 | 2 | 1 | 2 | 0 | 0 | $\frac{1}{p_0 p_1 p_2}$ | 1 |
| 3 | 3 | 1 | 0 | 0 | 0 | $\frac{1}{p_0 p_1 p_2}$ | 1 |
| 4 | 0 | 0 | 0 | 0 | $\frac{1}{p_0}$ | $\frac{1}{p_0}$ | 1 |
| 4 | 1 | 0 | 1 | 0 | $\frac{1}{p_0 p_1}$ | $\frac{1}{p_1 p_1}$ | 1 |
| 4 | 2 | 0 | 2 | 0 | $\frac{1}{p_0 p_1 p_2}$ | 0 | 1 |
| 4 | 3 | 1 | 3 | 0 | 0 | 0 | $\frac{1}{p_3}$ |
| 5 | 0 | 0 | 0 | 0 | $\frac{1}{p_0}$ | $\frac{1}{p_0}$ | 1 |
| 5 | 1 | 0 | 1 | 1 | $\frac{1}{p_0 p_1}$ | $\frac{1}{p_0 p_1}$ | 1 |
| 5 | 2 | 0 | 2 | 1 | 0 | 0 | 0 |
| 5 | 3 | 0 | 3 | 1 | 0 | 0 | 0 |

### A.9 Adjusting trial outcomes based on biology

Sometimes there is strong biological theory or evidence about the postexposure window in which vaccination is likely to be successful, for instance, when data from postvaccination serological assessments of antibody responses suggests meaningful change in immune responses occurs only after 7 days. In this case, there may be interest in restricting the time frame in which infection events count against vaccination. In a trial, this may be handled by re-defining the outcome such that only cases which occur more than 2 days post randomization are counted as events in both arms. Cases that occur prior to this are not counted in either trial arm. This is how outcomes were defined, for instance, in many of the trials of SARS-CoV-2 vaccines.

In observational emulations, we can similarly re-define vaccination outcomes based on biology, however we have to be careful to ensure that the new definitions are applied fairly across vaccination groups. In traditional analyses, bias can occur when all unvaccinated cases are counted from day zero but vaccinated cases are counted from the day of vaccination. This is fixed when using any of the target trial designs described previously because time zero is properly aligned in both groups. With a clear time zero in each nested trial or cloned regime, we can then unambiguously define delayed outcomes for all vaccination strategies. For instance, using the notation introduced previously, we define trial outcome *Y* ^*\**^ = *I*(*D*_Δ_ = 1, *D*_*δ*_ = 0) where *δ* here is the number of days after randomization in which cases do not count against the vaccine or no vaccine groups.

### A.10 Effect measures for vaccine effectiveness

In the main text, we defined vaccine effectiveness in terms of the cumulative incidence of symptoms or disease over the follow up period, e.g.

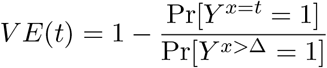

comparing vaccination regimes vaccinated on day *t* and never vaccinated over follow up. However, it is also common in the literature to see vaccine effectiveness defined instead in terms of hazards, e.g.

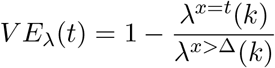

where *λ*^*x*^(*k*) is the (average) hazard rate over the follow up period, e.g.

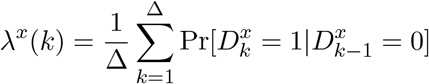

In the applied literature, these are sometimes used interchangeably even though they will rarely coincide, e.g. they will not coincide when hazard rates are nonconstant or heterogeneous or non-proportional. In the causal literature, there is a preference against causal hazard ratios particularly when they are time-varying (as they almost certainly are in practice) as they condition on survival and therefore introduce possible selection bias by construction.

However, previous work [5] has shown that patterns in *V E*(*t*) and *V E*_*λ*_(*t*) could, in some circumstances, help elucidate the mechanism of action of a particular vaccine, for instance to help distinguish whether a vaccine produces “all-or-none” or “leaky” protection against infection.

### A.11 Determining maximum postexposure vaccination delay

When setting guidelines for postexposure vaccination, a common problem is determining the maximum vaccination delay before effectiveness falls below a certain cost-benefit threshold. This quantity is important both for policymakers communicating with high risk groups and the broader public about what to do in the event of an exposure as well as to help practitioners determine whether vaccination is still indicated upon presentation. Absent clear biology or immune response data, it can be difficult to determine empirically even when postexposure trials are possible as trial participants are generally only assigned to vaccine or no vaccine/placebo not to a specific day to be vaccinated. In this section, we suggest a methods for estimating the maximum delay based on a pre-specified minimum effectiveness bound. In principle, these methods could be applied either in a randomized trial where the day of vaccination is not strictly controlled or in an observational emulation.

Suppose *u*(*Y* ^*g*^) is a utility function quantifying the health benefits of vaccination strategy *g* and *V* is a subset of covariates defining subpopulations of interest, such as certain high risk exposure groups, then the conditional mean

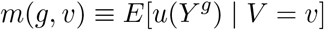

is the expected utility under a hypothetical policy in which everyone in the subpopulation receives vaccination strategy *g*. Comparing the expected utility *m*(*g, v*) for across different regimes *g ∈*𝒢 quantifies the counterfactual benefits of vaccination under different delays.

To determine the optimal guidance regarding postexposure delays, we might consider two distinct classes of regimes and subpopulations (although others are certainly possible):

- A class of day zero regimes in which everyone is vaccinated after a delay of *x*^*\**^ days, i.e. *g*(*x*) = *x*^*\**^ and *V* = 1 for everyone.
- A class of regimes in which all of those remaining unvaccinated and symptom-free on day *x*^*\**^ are vaccinated, i.e. *g*(*x*) = *x*^*\**^ and *V* = *I*(*X > x*^*\**^, *T > x*^*\**^).

Each answers a slightly different question and may be relevant under different circumstances. The second is more relevant for practitioners counseling patients who present symptom-free on their options after exposure, while the first is more relevant for public health guidance telling those currently unexposed how quickly they need to get to a clinic after exposure.

Our goal is then to find the maximum value of *x*^*\**^ in which the utility in subpopulations of interest remains above some minimum viable threshold, i.e.

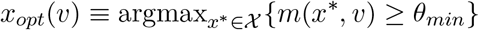

To take a simple example, consider the case that *m*(*x*^*\**^, *v*) is just the vaccine effectiveness (i.e. there are no other costs or benefits) in which case we want to solve

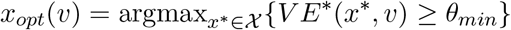

where

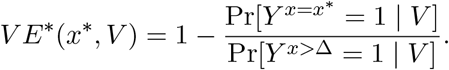

In this case, *x*_*opt*_(*v*) is the maximum delay before vaccine effectiveness falls below some minimum threshold.

To determine *x*_*opt*_(*v*) for the first class of regimes, one approach would be to calculate 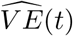 from the day zero design separately for each regime as *V E*(*t*) = *V E*^*\**^(*t*, 1) and then determine the maximum value of *t* where 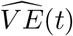 remains above the threshold. However, we can also increase efficiency by pooling across the cloned strategies and fitting a model with a flexible function of the delay regime such as that in **??**. We can then estimate the 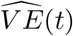 curve either from estimated hazard ratios or from standardized cumulative incidence curves depending on effect measure of interest and using inverse probability of censoring weights to adjust for nonadherence among unvaccinated where applicable.

Alternatively, to determine *x*_*opt*_(*v*) for the second class of regimes, we could use the stratified estimates 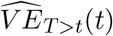 from each of the nested daily trial emulations as *V E*_*T >t*_(*t*) = *V E*^*\**^(*t, I*(*X > t, T > t*)) and then determine the maximum value of *t* where 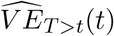 remains above the threshold. However, this assumes we observe sufficient numbers of individuals being vaccinated on each day to obtain reliable estimates. In practice, we might prefer to increase efficiency by pooling across trials and fitting a model with a flexible function of vaccination timing such as that in **??**. We can then estimate the 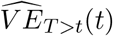 curve either from estimated hazard ratios or from standardized cumulative incidence curves depending on effect measure of interest and using inverse probability of censoring weights to adjust for nonadherence among unvaccinated where applicable.

### A.12 Simulation

To demonstrate the benefits of the target trial approach, we simulated data from hypothetical observational study under a known data generation process in which there is an overlap between vaccination timing and the timing of symptom onset. We used this setup to compare explicit emulation of a target trial with a few common estimation strategies drawn from the literature. In all simulations, we drew datasets of size 1000, estimated the *V E* under each estimation strategy, and repeated the across 1000 Monte Carlo samples to calculate the bias and efficiency.

#### A.12.1 Data generating mechanism

We simulated postexposure vaccination by first drawing a vaccine “assignment” indicator, *Z*, from a Bernoulli distribution with probability 0.5 and then drawing a postexposure delay, *X*^*\**^, from a Poisson distribution with a mean of 5 days. In an observational study, we assume that true assignment is unknown and therefore we only observe vaccination times among the vaccinated, i.e. *X* = *ZX*^*\**^. We then simulated symptom onset over the 21 days of follow up after exposure based on the discrete time hazard model

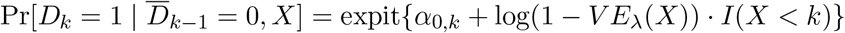

for *k* in {0, …, 21} where *Y* = *D*_21_ and the baseline hazard *α*_0,*k*_ was defined such that there is a 50% probability of symptoms given exposure among unvaccinated and onset times among cases had a Log-Normal distribution with parameters chosen based on previous estimates of the incubation period for mpox [6]. We assumed vaccination reduces probability of symptoms but does not affect onset timing and only works if administered prior to onset. For those with simulated vaccination times that occur after symptom onset we assumed 25% still receive the vaccine, while vaccination time was censored for the remaining. We generated data under three scenarios for vaccine effectiveness:

- *scenario 1:* the null case that vaccination is completely ineffective, i.e.

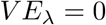
- *scenario 2:* vaccination reduces hazard of symptom onset by a constant of 40% (corresponding to 21-day VE of 31.6% based on cumulative incidence), i.e.

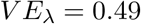
- *scenario 3:* a more realistic scenario in which effectiveness is a function of postexposure timing, i.e.

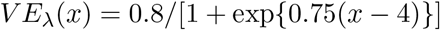

The full data generation process may be written as:

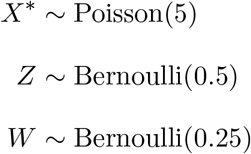

for *k* ∈ {1, …, 21} : *D*_*k*_ *∼* Bernoulli(expit{*α*_0,*k*_ + log(1 *− V E*_*λ*_(*X*^*\**^))· *Z* · *I*(*X*^*\**^ *< k*)*}}*)

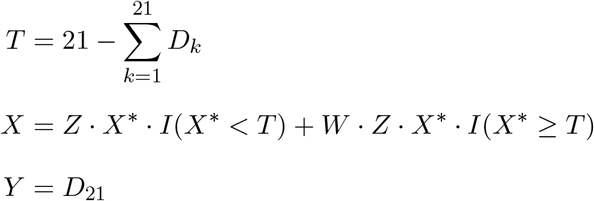

where

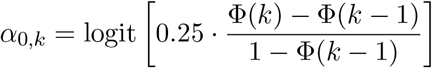

and Φ is the cumulative distribution function for a log-normal distribution with log mean of 2.1 and log standard deviation of 0.59.

Figure A5 shows the overlap in the distribution of vaccination times and disease onset times when *V E* = 0. Note that under this process, there is no structural source of confounding, i.e. vaccination status and timing is random with respect to symptom onset. Rather bias comes from the true “assignment” being unknown to the investigator.

##### A.12.2 Target and estimation: V E based on the cumulative incidence

In the first set of simulations, our target parameter was the vaccine effectiveness based on the relative reduction in cumulative incidence, i.e.

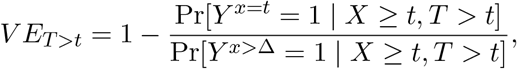

which is the same as the estimand defined for an ideal trial based on a fixed-enrollment period design. We compared three different estimation strategies:

1. *naïve, leave* - a simple comparison of the “ever vaccinated” and “never vaccinated” using the relative risk regression model Pr[*Y* = 1 | *X*] = exp{*β*_0_ + *β*_1_*I*(*X <* 21)} and vaccine effectiveness is estimated as 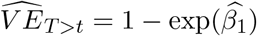.
2. *naïve, move* - those who receive vaccine after developing symptoms are re-classified as “unvaccinated”, i.e. we use the relative risk regression model Pr[*Y* = 1 | *X*] = exp{*β*_0_ +*β*_1_*I*(*X < T*)} where *I*(*X < T*) implies only those who receive vaccine prior to symptom onset are “vaccinated” and vaccine effectiveness is estimated as 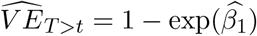 as before.
3. *target trial* - we emulate a sequence of nested daily trials by taking those who are symptom free and unvaccinated prior to start and compare those are vaccinated on that day to those who are not. In each trial, we censor the unvaccinated when they become vaccinated and use inverse-probability of censoring weights to account for informative censoring. These nested trials are combined and vaccine effectiveness is estimated using standardized cumulative incidence curves from a pooled logistic regression and standard errors are estimated using cluster-robust variance estimator.

The first two are strategies that we have seen used in observational studies of post-exposure vaccination and the last is the one proposed in this paper.

#### A.12.3 Target and estimation: V E based on the hazard

In another set of simulations, we also considered targeting the vaccine effectiveness based on the (average) hazard rather than the cumulative incidence of symptom onset, i.e.

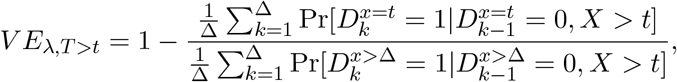

We compared four different estimation strategies:

1. *naïve, leave* - similar to above however we estimate incidence rates rather than cumulative incidence through poisson regression Pr[*Y* = 1 | *X*] = exp{*β*_0_ + *β*_1_*I*(*X <* 21)} with offset log(*T*) and vaccine effectiveness is estimated as 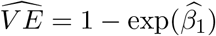.
2. *naïve, move* - those who receive vaccine after developing symptoms are re-classified as “unvaccinated”, i.e. we use the poisson regression model Pr[*Y* = 1 | *X*] = exp{*β*_0_ + *β*_1_*I*(*X < T*)} with offset log(*T*) and *I*(*X < T*) implies only those who receive vaccine prior to symptom onset are “vaccinated” and vaccine effectiveness is estimated as 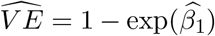 as before.
3. *time-varying cox* - use a time-varying cox model *λ*(*t*|*X*) = *λ*_0_(*t*) exp{*β*_1_*I*(*X ≥ t*)} in which follow up time is split for vaccinated participants at the time of vaccination. Prior to this their person time is classified as unvaccinated and effectiveness is estimated as 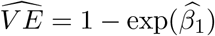.
4. *target trial* - same as previous, except we estimate vaccine effectiveness as one minus the exponentiated coefficient from the pooled logistic regression model rather than from standardized cumulative incidence curves.

#### A.12.4 Target and estimation: comparing trial targets

In a final set of simulations, we consider the same data generation mechanism but targeting different hypothetical trials to show the flexibility of our approach to answer different research questions. Specifically, we consider:

- *fixed-enrollment:* a trial in which participants who are symptom free are enrolled within 7-days postexposure and randomized to either receive a vaccine or no vaccine on the day of enrollment. We emulate this in the simulated observational data as a sequence of nested daily trials as discussed in A.6.1. In each trial, we censor the unvaccinated when they become vaccinated and use inverse-probability of censoring weights to account for informative censoring. These nested trials are combined and vaccine effectiveness is estimated using standardized cumulative incidence curves from a pooled logistic regression with *V E* by day modeled using flexible cubic splines and standard errors are estimated using cluster-robust variance estimator.
- *day-zero:* a trial in which participants are randomized on the day that they were exposed to vaccine or no vaccine and then also randomized a day between 0 and 6 to receive the vaccine. We emulate this using the clone-censor-weight approach described in A.6.2. We censor clones when they deviate from their assigned regime and use inverse-probability of censoring weights to account for informative censoring. These nested trials are combined and vaccine effectiveness is estimated using standardized cumulative incidence curves from a pooled logistic regression with *V E* across delay regimes modeled using flexible cubic splines and standard errors are estimated using cluster-robust variance estimator.
- *grace period* a trial in which participants are randomized on the day that they were exposed to vaccine or no vaccine with those assigned to vaccine allowed a 7 day grace period in which to receive a vaccine. We emulate this using the clone-censor-weight approach described in A.6.3. We censor clones when they deviate from their assigned regime and use inverse-probability of censoring weights to account for informative censoring. These nested trials are combined and vaccine effectiveness is estimated using standardized cumulative incidence curves from a pooled logistic regression and standard errors are estimated using cluster-robust variance estimator.

##### Simulation results

In Figure A6 (and Table A5), we compare estimates of *V E* based on the cumulative incidence of symptom onset to the truth for scenarios 1 and 2. Under the null, the naïve approaches are upwardly biased due to immortal time bias (i.e. by definition vaccinated have to survive long enough to be vaccinated while unvaccinated are at risk at all time points), while the target trial approaches yield valid estimates. This persists in scenario 2 where *V E* = 31.6%, although the relative bias of the first approach is somewhat offset by the fact that those vaccinated after developing symptoms are included with vaccinated. In scenario 3, where vaccine effectiveness varies with postexposure timing, the naïve approaches still produce biased estimates, with larger bias for greater postexposure delays. The target trial approach yields unbiased estimates of vaccine effectiveness at all time points (Figure A7).

Another common approach to account for immortal time is to split follow up at the time of vaccination among the vaccinated and use a time-varying specification of the Cox proportional hazards model to estimate *V E*. In Figure A8 (and Table A6), we show this approach also yields unbiased estimates of postexposure vaccine effectiveness when evaluated using one minus the hazard ratio rather than cumulative incidence (the latter could, in theory at least, be obtained by combining with a suitable estimator of the baseline hazard, but this is uncommon). However, in practice, this method imposes restrictions on appropriate adjustment for time-varying confounding that is almost certainly present in most real world applications. In contrast, the naïve methods based that estimate the average hazard or incidence rate are biased, again due to immortal time bias.

To explore the extent to which the overlap between vaccination and symptom onset drives our results, we also evaluated how performance varies with the degree of overlap. Specifically, we repeated scenario 2 (*V E* = 31.6%) for VE based on the cumulative incidence and varied the mean of the log-normal distribution used to generate the symptom onset times. Here, larger mean onset times correspond to later symptom onset and thus less overlap. In Figure A9, we show that the bias of the naïve approaches increases as the mean onset time gets shorter while both the target trial and time-varying Cox approaches remain unbiased. This suggests that the target trial approach may be particularly useful in settings with high overlap between vaccination and symptom onset or those in which the majority of cases occur prior to vaccine being administered.

Finally, in Figure A10 we show the versatility of the target trial framework for targeting different trial designs/strategies. Using the modified emulations discussed above and in section A.6, the target trial approach produces unbiased estimates of a trial with a 7-day fixed enrollment period, a trial where participants are cross-randomized on day zero postexposure to vaccination and a delay of between 0 and 6 days, and a trial where participants are randomized to vaccine or no vaccine on day zero postexposure but given a 7-day grace period in which to receive vaccination.

**Table A5:** Simulation results for scenarios 1 and 2 when estimating VE using the risk ratio.

| Estimator | Mean | Bias | SD | RMSE |
| --- | --- | --- | --- | --- |
| *Scenario 1: $VE = 0\%$* |  |  |  |  |
| naive, leave | 0.090 | 0.090 | 0.059 | 0.108 |
| naive, move | 0.120 | 0.120 | 0.058 | 0.133 |
| target trial | -0.004 | -0.004 | 0.067 | 0.067 |
| *Scenario 2: $VE = 31.6\%$* |  |  |  |  |
| naive, leave | 0.377 | 0.060 | 0.050 | 0.078 |
| naive, move | 0.407 | 0.091 | 0.049 | 0.103 |
| target trial | 0.317 | 0.000 | 0.057 | 0.057 |

**Table A6:** Simulation results for scenarios 1 and 2 when estimating VE using hazard ratio

| Estimator | Mean | Bias | SD | RMSE |
| --- | --- | --- | --- | --- |
| *Scenario 1: $VE = 0\%$* |  |  |  |  |
| naive, leave | 0.146 | 0.146 | 0.072 | 0.163 |
| naive, move | 0.192 | 0.192 | 0.068 | 0.203 |
| time-varying cox | -0.007 | -0.007 | 0.095 | 0.096 |
| target trial | -0.008 | -0.008 | 0.098 | 0.099 |
| *Scenario 2: $VE = 40\%$* |  |  |  |  |
| naive, leave | 0.456 | 0.056 | 0.052 | 0.077 |
| naive, move | 0.494 | 0.094 | 0.050 | 0.107 |
| time-varying cox | 0.395 | -0.005 | 0.065 | 0.066 |
| target trial | 0.400 | 0.000 | 0.067 | 0.067 |

**Figure A5:**
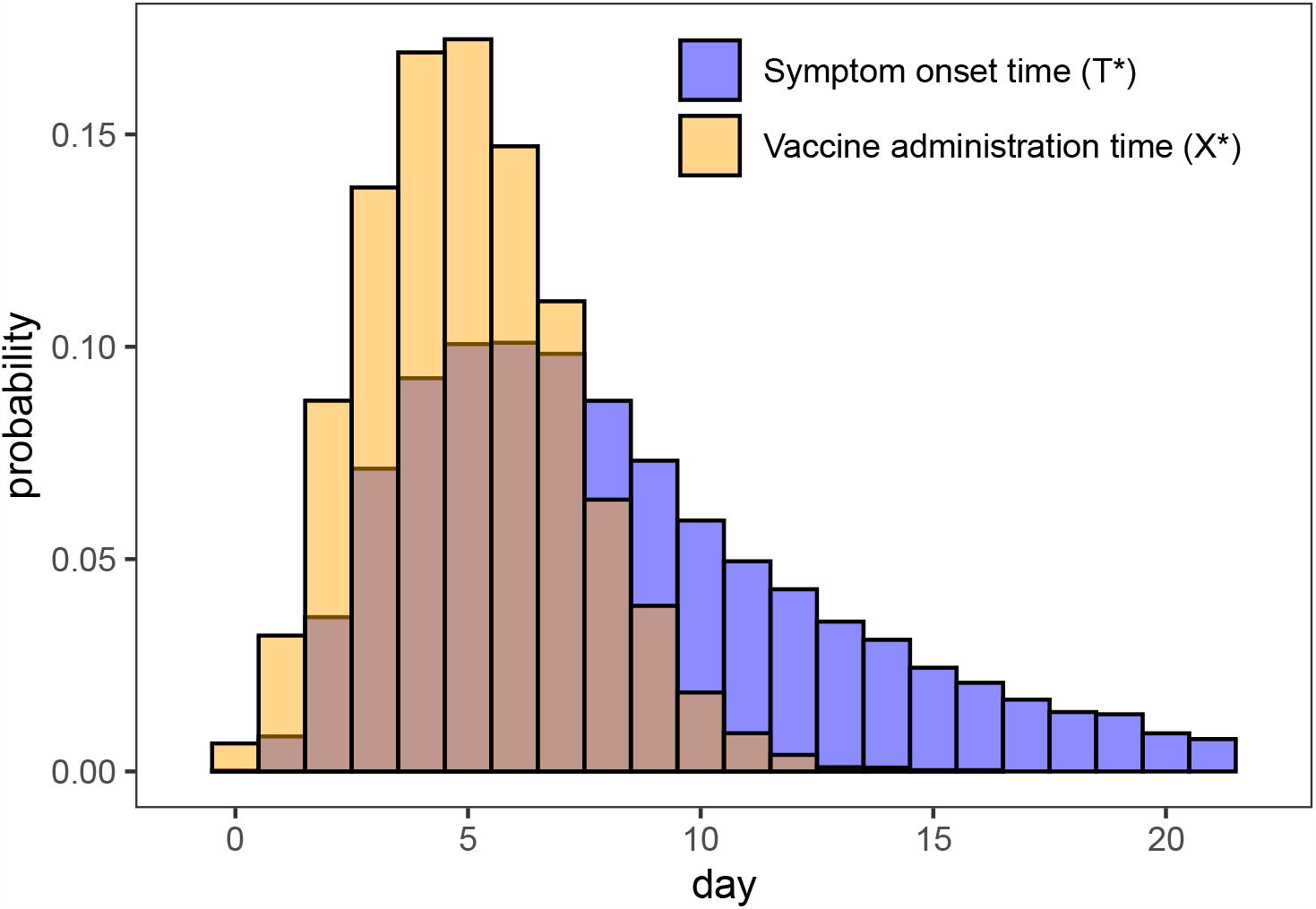
Distribution of simulated vaccination times (*X*^*\**^) among vaccinated and symptom onset times (*T* ^*\**^) among cases when *V E* = 0 over the 21 days of follow up showing the degree of overlap.

**Figure A6:**
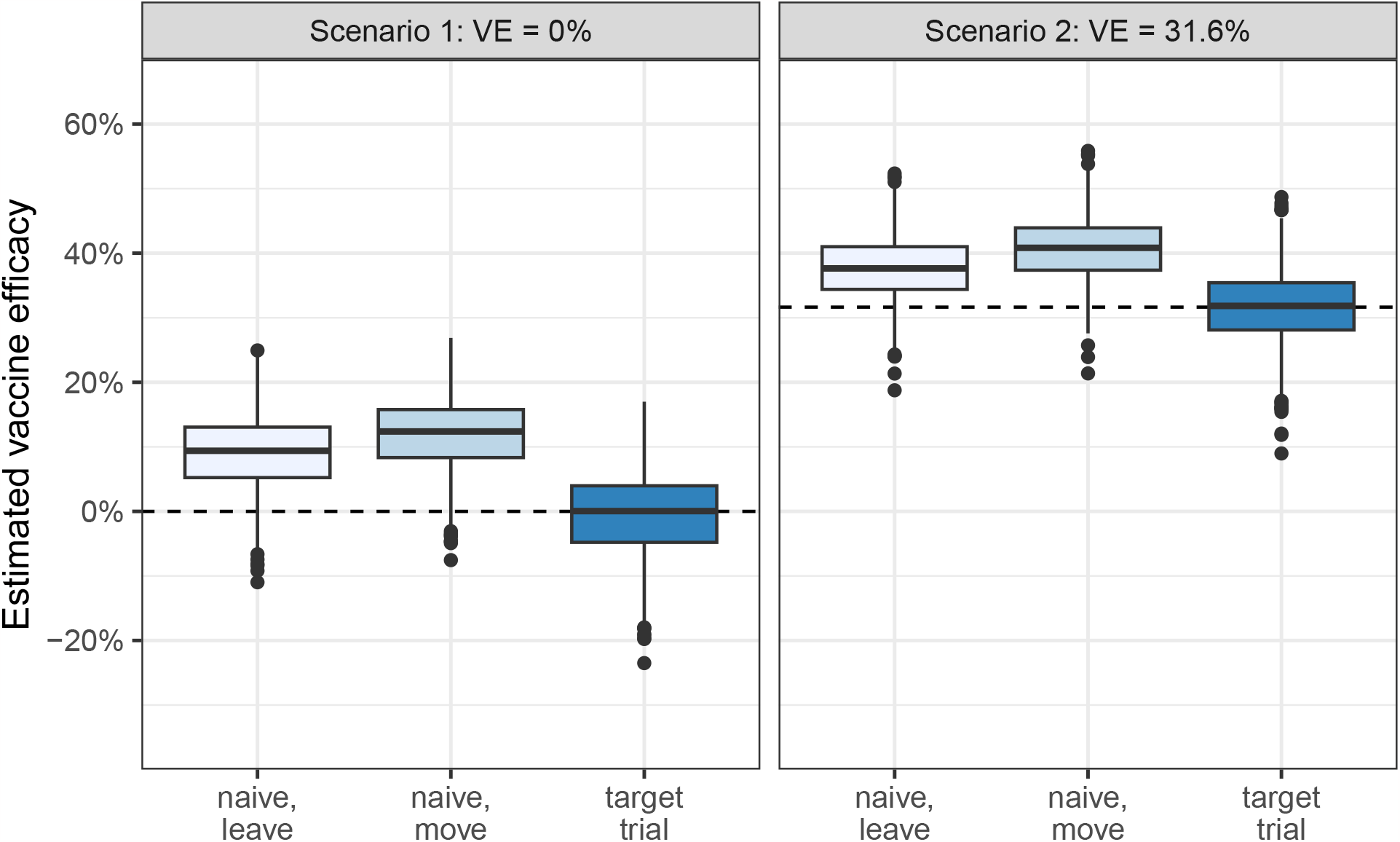
Simulated *V E* estimates compared to the truth for the three estimation strategies described in section 5. Based on 1000 monte carlo simulations. Dashed line shows true value in each scenario.

**Figure A7:**
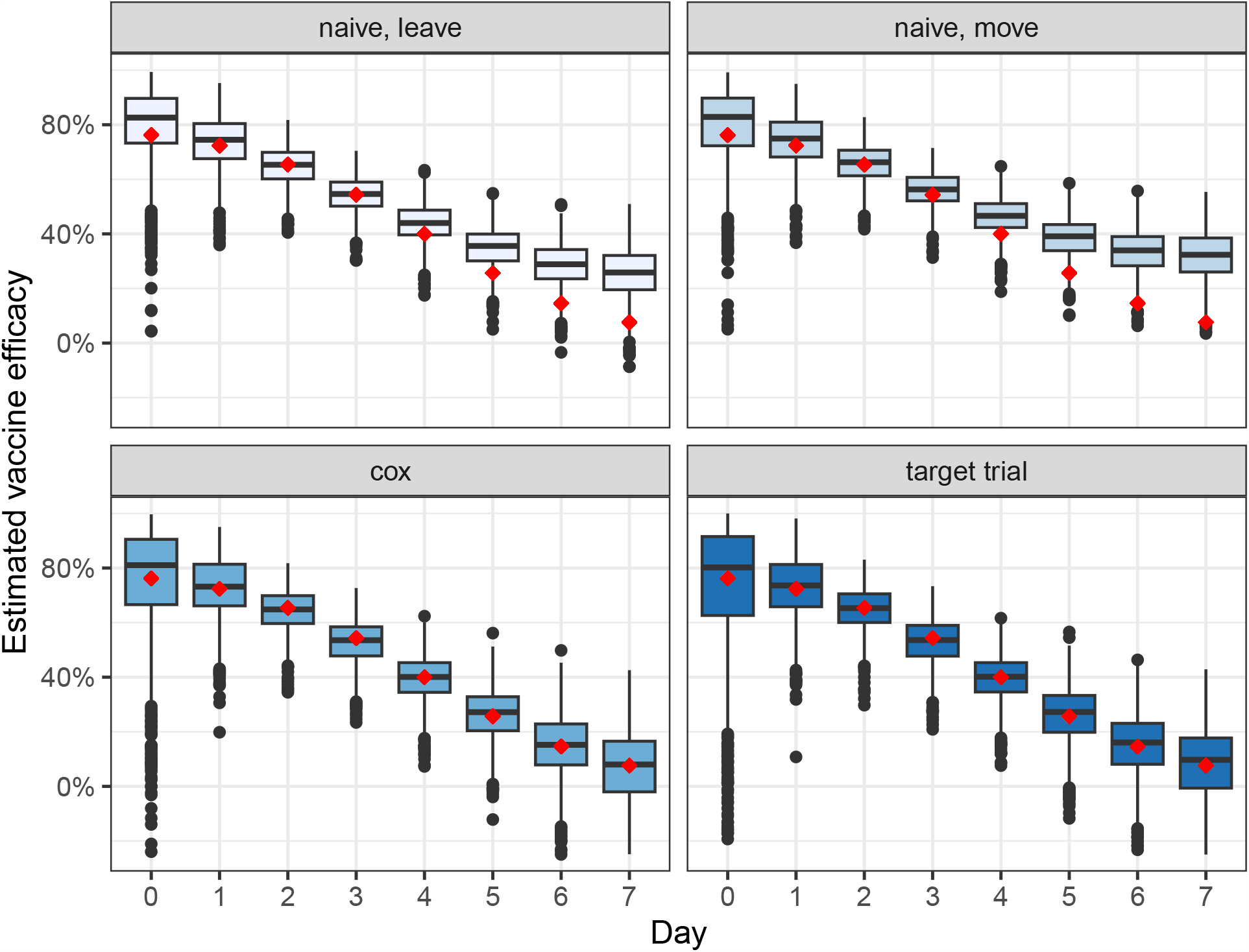
Comparison of estimators under when vaccine effectiveness varies by postexposure administration time.

**Figure A8:**
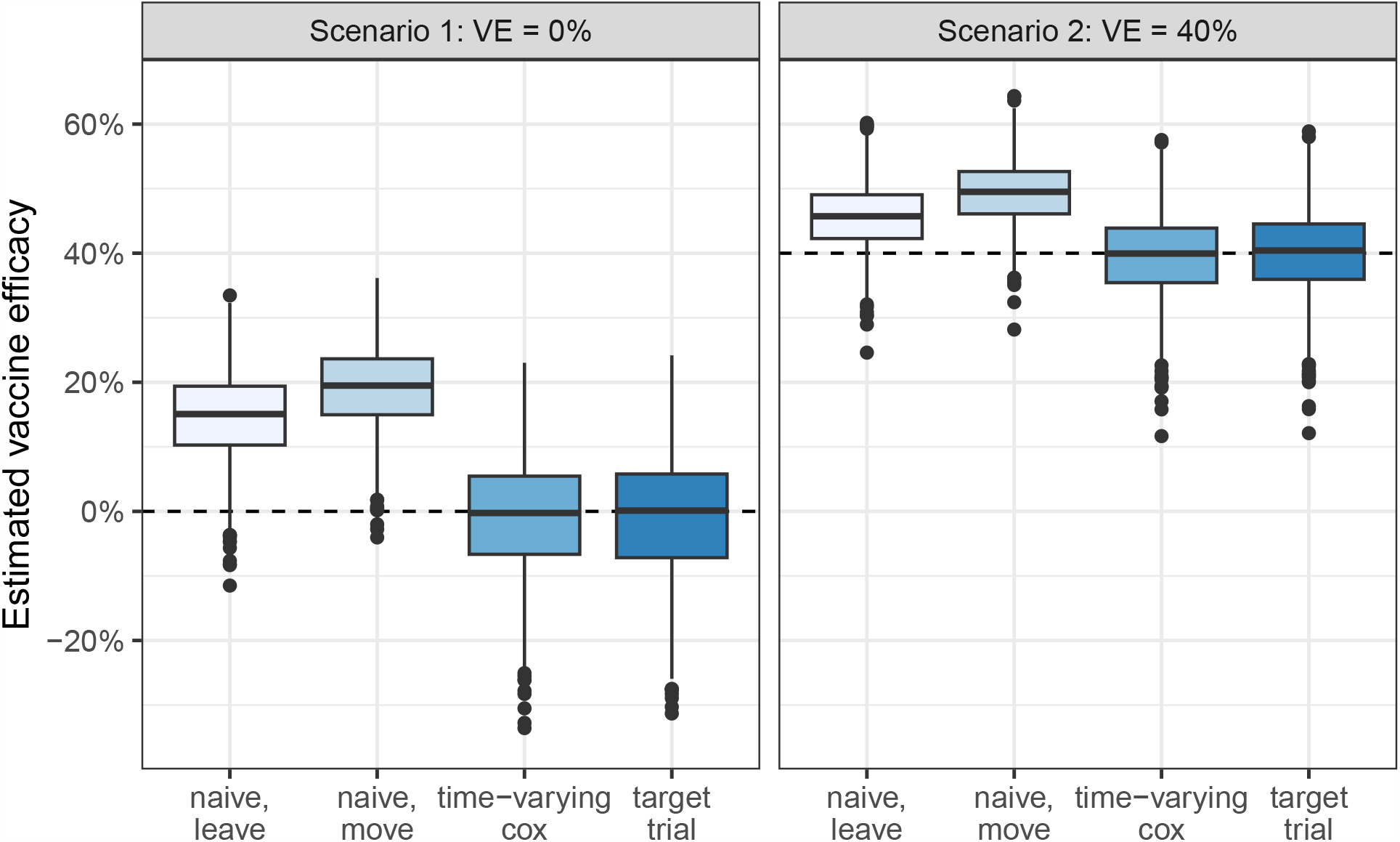
Comparison of estimators when calculating vaccine effectiveness using the hazard ratio instead of the risk ratio.

**Figure A9:**
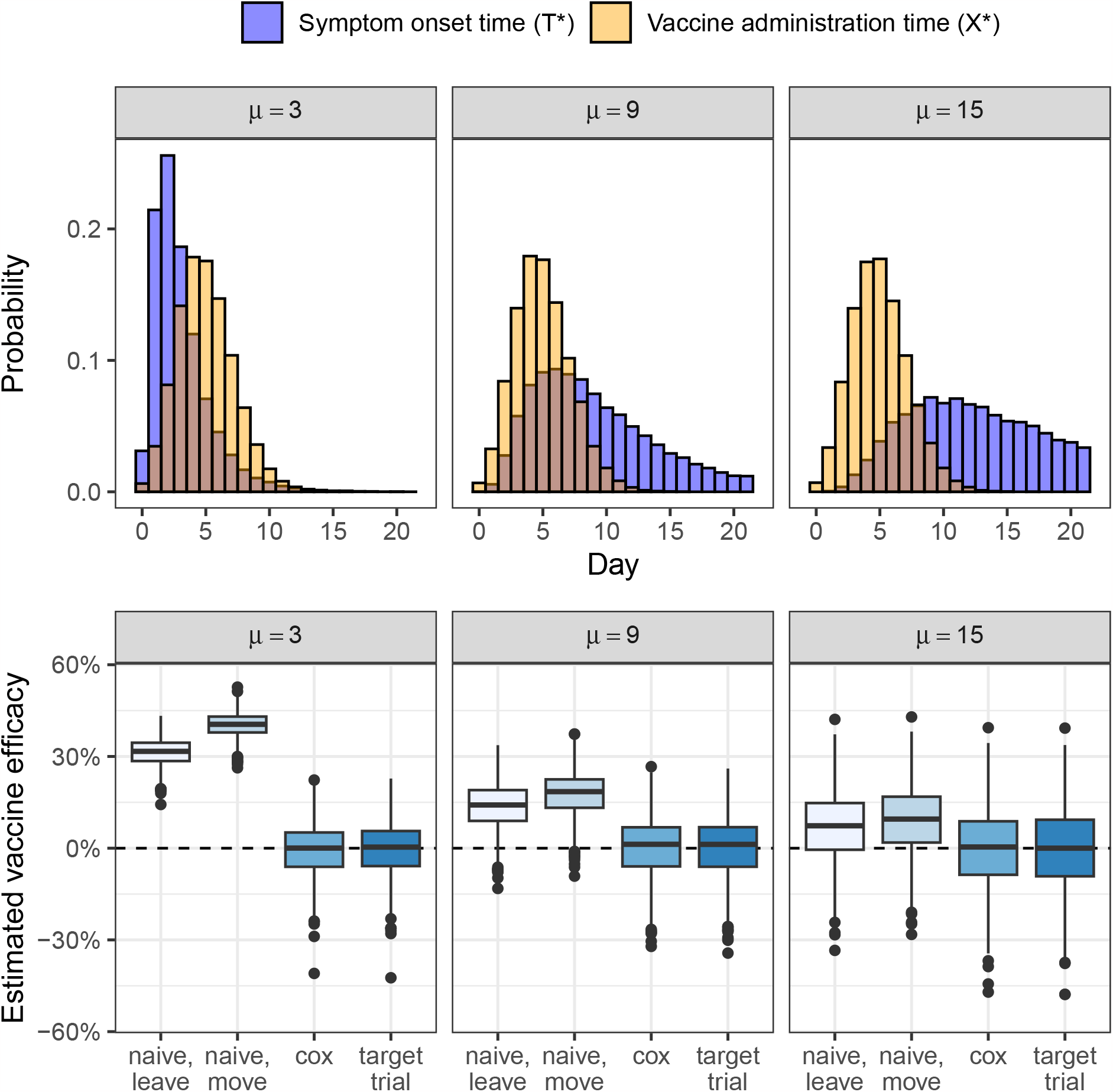
Bias of naïve methods varies with degree of overlap between vaccination delays and symptom onset times.

**Figure A10:**
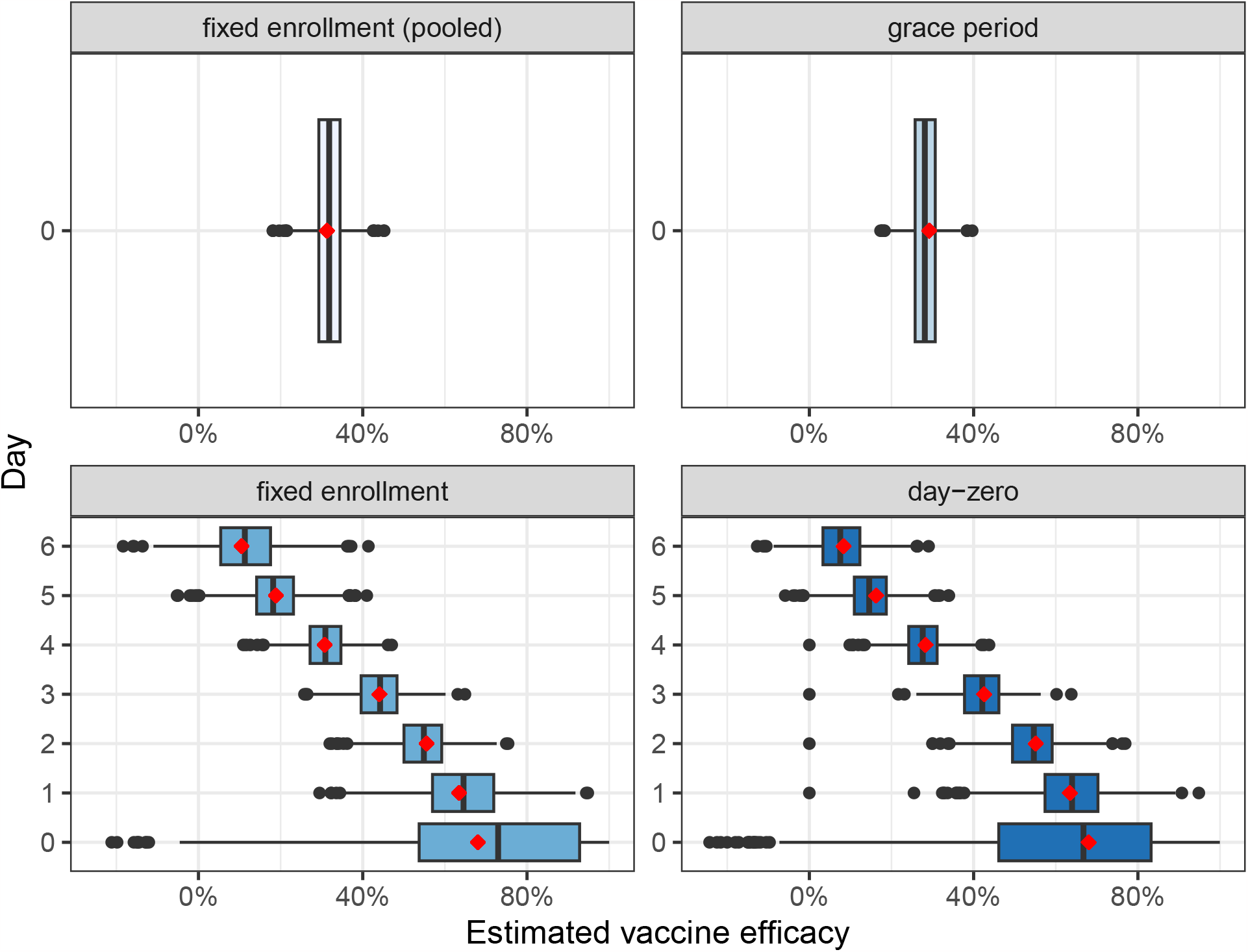
Simulated *V E* estimates compared to the truth for four different target trial designs described in section 5. Based on 1000 monte carlo simulations. Red dots shows true value in each scenario.

## Notes

### Competing Interest Statement

In the past 24 months, ML has received grant support from Pfizer and consulting income from Janssen. He is Senior Advisor to the CDC's Center for Forecasting and Outbreak Analytics, but this contribution is in his academic role and does not necessarily express the views of any government entity.

### Funding Statement

This work was funded by Morris-Singer Fund's gift to the Center for Communicable Disease Dynamics

### Summary of Updates

Fixed typos in Table A4.

